# Evidence of ethnic variations in the relationships between routinely recorded clinical factors and T2D: a systematic review and meta-analysis

**DOI:** 10.1101/2025.04.07.25325365

**Authors:** B Orazumbekoba, T Hamdani, S Hodgson, M Samuel, D Stow, M Spreckley, S Finer, M K Siddiqui, R Mathur

## Abstract

**Background:** Evidence on ethnic differences in factors associated with type 2 diabetes (T2D) is mixed. We aimed to systematically review evidence on ethnic variations in the relationships between routinely recorded demographic and clinical factors and T2D.

**Methods:** We searched Medline Complete and Embase for observational studies published between 1990 and 2023 investigating ethnic differences in factors routinely recorded in clinical encounters associated with T2D. We used random and fixed-effects meta-analysis to quantitatively summarise effect sizes across studies where possible. Risk of bias and study quality were assessed using the Newcastle-Ottawa Scale and Joanna Briggs Institute tool. PROSPERO registration: CRD42023394148.

**Findings:** Searches identified 10 694 studies, of which, 54 (n=10 332 949 individuals) were eligible for inclusion, including 12 suitable for meta-analysis. Included studies reported ethnic differences in age at T2D diagnosis, anthropometric measures, and factors associated with women’s health. Compared to individuals of White ethnicity, people of diverse ethnic backgrounds had 2-4-fold higher incidence and prevalence of T2D and younger age of onset. Waist-to-hip ratio (WHR) was a better discriminator of T2D across all ethnic groups compared to body mass index (BMI). While the association between overweight/obese BMI and T2D was strongest for people of White ethnicity (OR 4.85 CI 3.53-6.68) followed by Black (OR 3.27 CI 2.48-4.30) and East Asian ethnicities (OR 3.06 CI 2.29-4.16), the association between WHR and T2D was strongest for people of Black (OR 2.74, CI 2.22-3.39) than for White ethnicities (OR 2.51, CI 2.30-2.74). Included studies highlighted the emerging importance of women-health-associated factors such as index of parity, birth weight and breastfeeding, especially among women of diverse ethnicities.

**Conclusion:** Ratio measures of central adiposity may better identify T2D in ethnically diverse populations than measures of overall adiposity. Sex-specific factors must be considered when assessing T2D risk.

**Funding:** Wellcome Trust Grant GPPG1K9R/218584/Z/19/Z.

## Introduction

Type 2 diabetes (T2D) is a global public health problem, which, as of 2021, affects over 537 million people worldwide (1). This number is expected to increase to 783 million by 2045 (1) along with associated healthcare expenditure (2). The prevalence and incidence of T2D varies by ethnicity: (3, 4) people of Asian ethnicity (3, 5, 6) especially South Asian (3, 7), Black and Arab ethnicities(8), and individuals of other ethnic backgrounds (5, 6) have at least a two to fourfold risk of developing T2D compared to people of White ethnicity. People of these ethnicities are also at a greater risk of developing diabetes-related micro- and macro-vascular complications.(9)

Previous research suggests the pathophysiology of T2D differs across ethnicities; While insulin resistance is not a major driver of diabetes in White populations, it plays an important role in other ethnic groups, related to ß-cell dysfunction in people of South Asian ethnicity and lower hepatic insulin clearance in people of Black ethnicity (10, 11). There is a growing body of literature indicating people of the above-mentioned ethnic backgrounds develop T2D at younger ages compared to people of White ethnicity (8, 12, 13). Along with this, the proportion of individuals who develop T2D within normal body weight ranges (BMI 18.5-24.9 kg/m2) is higher among individuals of different ethnic backgrounds compared to people of White ethnicity (8, 14). Current guidelines for the prevention of T2D are based on evidence largely derived from White European populations and are limited to decreasing BMI for people with overweight and obesity and for certain ethnic groups, identifying diabetes risk at earlier ages (15, 16). However, the differing causal mechanisms for T2D are still poorly understood and current guidelines may be inadequate for managing T2D and its longer-term sequelae in diverse ethnic populations. While several systematic reviews and meta-analyses have described key factors associated with T2D (17–19), none have yet focussed on systematically comparing these across ethnic groups.

To inform clinical risk prediction and facilitate the translation of routinely available health data into practice, we systematically reviewed published evidence on ethnic differences in the relationships between demographic and clinical factors routinely captured in clinical care settings and type 2 diabetes.

## Methods

We registered this systematic review with PROSPERO (registration number CRD42023394148). This study was funded by the Wellcome Trust Grant, reference 218584/Z/19/Z. The funder of the study had no role in study design, data collection, data analysis, data interpretation, or writing of the report.

### Data sources and Search strategy

We searched Medline Complete and Embase databases according to the Preferred Reporting Items for Systematic Reviews and Meta-Analyses (PRISMA) guidelines. To ensure coverage of all potentially eligible studies, we prepared a list of T2D- and ethnicity-related Medical Subject Headings (MeSH) and keywords (**Table S1**).

### Selection process and Data extraction

Study eligibility was assessed by two independent reviewers (BO and TH). We retrieved and merged search results from Medline Complete and Embase databases using Endnote reference management software. We removed duplicates and exported a list of unique studies to Rayyan reference management software. (20) The screening stage was blinded to both reviewers. After screening, conflicting results were discussed with the research team, following which we assessed full texts for eligibility. We contacted the authors of one paper without full text available to request access.

We included studies that met the following eligibility criteria: Studies published in English between 1990-2023, studies with an observational design (cross-sectional, case-control, cohort studies) focusing on the association between routinely collected demographic and clinical factors and T2D; studies which compared two or more ethnic groups, and studies where the outcome was T2D. We excluded studies that only reported results for one ethnic group. By doing so we focussed our systematic review on studies with the explicit aim of comparing effects across ethnicities. We excluded studies where the outcome was another type of diabetes (e.g. Type 1 diabetes, gestational diabetes), as well as prediabetes, impaired fasting glucose, impaired glucose tolerance and insulin resistance. We also excluded studies restricted to populations with specific health conditions or receiving specific treatments (e.g. people with serious mental illness, cancer, etc.); studies focusing on factors not routinely available in clinical records (genetics, lifestyle factors etc.) or studies with small sample per ethnic group, which could be underpowered to find clinically informative associations (total n<200 or fewer than 50 subjects per ethnic group).

### Risk of bias assessment

We used the Newcastle-Ottawa Scale (NOS) (21) and Joanna Briggs Institute tool (JBI) (22, 23) to assess the risk of bias. Two reviewers conducted the assessments independently and disagreements were discussed with a third senior reviewer. The risk of bias for each study was assigned as low, moderate or high risk.

### Data analysis

We conducted a narrative synthesis structured around the association between risk factors of interest and T2D across ethnicities and, if available, by age and sex. In studies reporting comparable associations, effect sizes and statistical significance were synthesised for each risk factor by ethnic group, as outlined by guidance on the conduct of narrative synthesis for systematic reviews (24). Due to a lack of standardised approaches to grouping ethnicities, we aggregated findings into broad ethnic groups, as detailed in **Table S2**. This categorization facilitated the summarization and reporting of more generalisable findings. For example, individuals of Pakistani, Bangladeshi and Indian ethnicities were groups as South Asian. However, if the effect of risk factors was compared across single ethnicities the ethnic categorization as reported by the original study was used.

Associations between routinely recorded clinical factors and T2D were meta-analysed and compared across ethnic groups when two or more studies reported on the same routinely collected factor(s). Given the heterogeneity in study designs and population characteristics (location, age groups, sex) random effect meta-analysis was used where more than two studies were included in meta-analysis, while fixed effects model meta-analysis was used where only two studies were eligible (25). The heterogeneity between studies was quantified by I^2^ statistics, which ranges between 0% and 100% - greater indicating a higher between-study variability (25). Where possible, we have provided sex-stratified meta-analysis estimates. Funnel plots were plotted to assess publication bias. Analysis was conducted using the *meta* package in R (26) and the *metan* package in Stata 15 (27).

## Results

The study selection process is represented in the PRISMA flow chart (**Fig. 1**). We identified 10 694 papers from two databases. After removing duplicates, 8 552 titles and abstracts were screened, of which 8 031 studies did not meet eligibility criteria. We reviewed the full texts of 521 articles and identified 59 eligible studies, including four from the reference lists of selected papers. Five risk factors were only reported in one study. Results from these studies are reported in the supplementary materials (**Table S3**). Due to women’s health having several factors identified in the same domain, these are discussed in the main text (pregnancy index, breastfeeding, birth weight, and age at menarche), leaving 54 eligible, full-text papers. We described and narratively summarized the 54 included studies; 12 studies were included in the meta-analysis of BMI (n=4 for OR, 2 for HR and 2 for RR, WHR (n=2) and family history of diabetes (FHD) (n=2).

**Figure 1.**
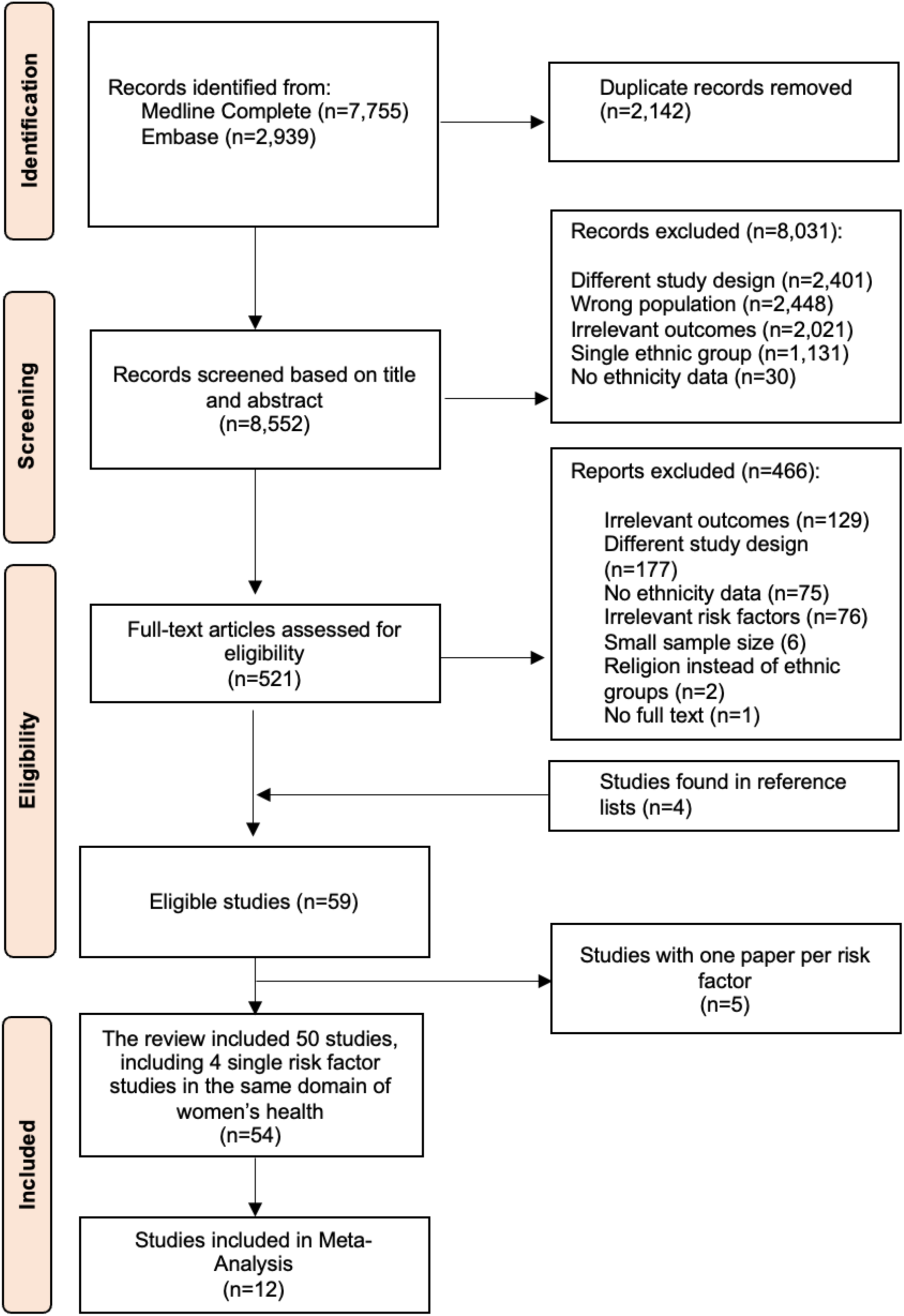
PRISMA flow chart

### Study characteristics and risk of bias

Out of 54 studies, there were 29 cross-sectional, 21 cohort and 4 case-control studies coming from 15 countries and several combined populations from different countries, with an overall number of participants of 10 337 613. The main ethnic groups included in the studies were South Asian, Southeast Asian, Black, East Asian, White, Hispanic and others **(Table S2**). Full study characteristics are shown in **Table 1**.

**Table 1.**
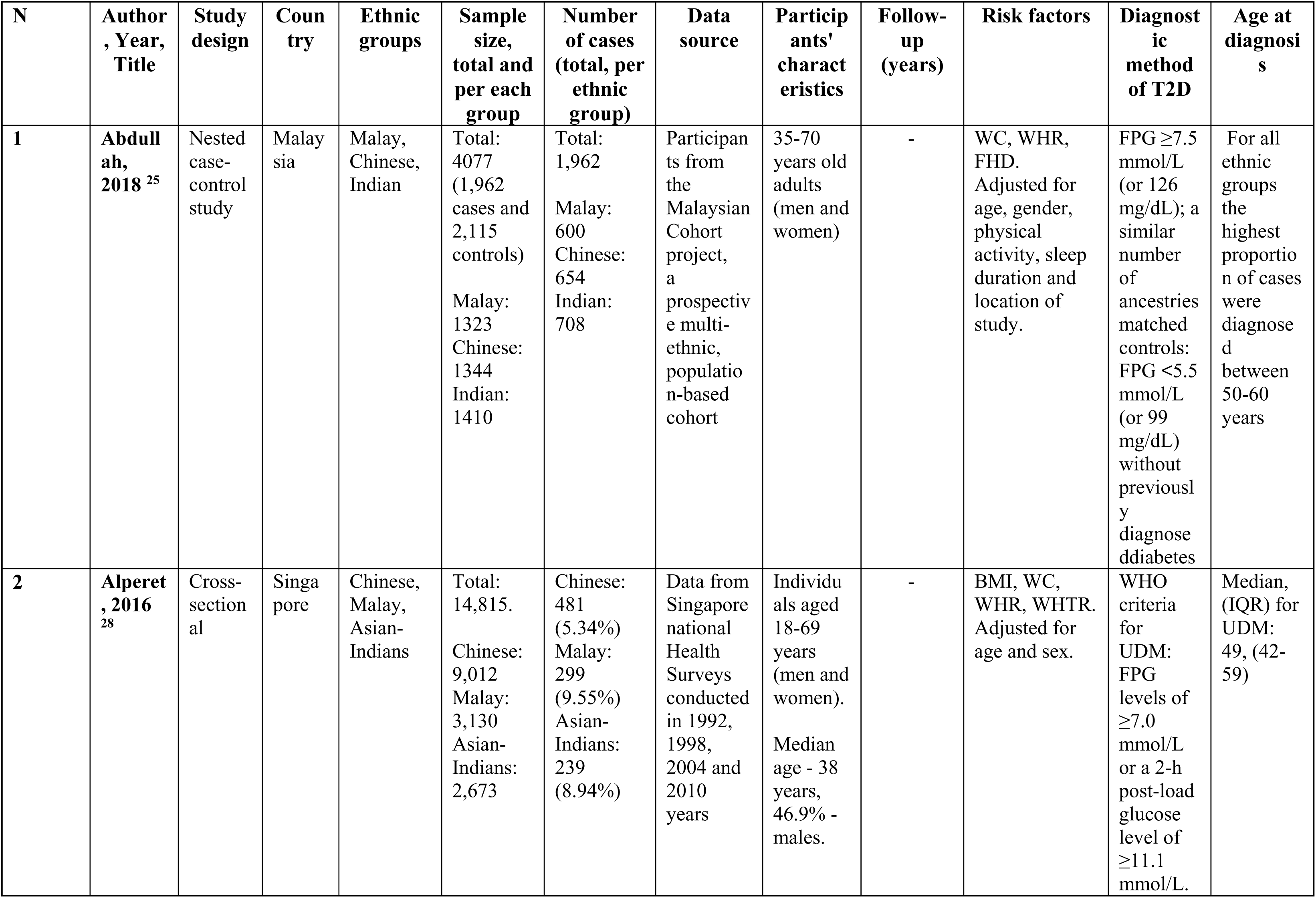
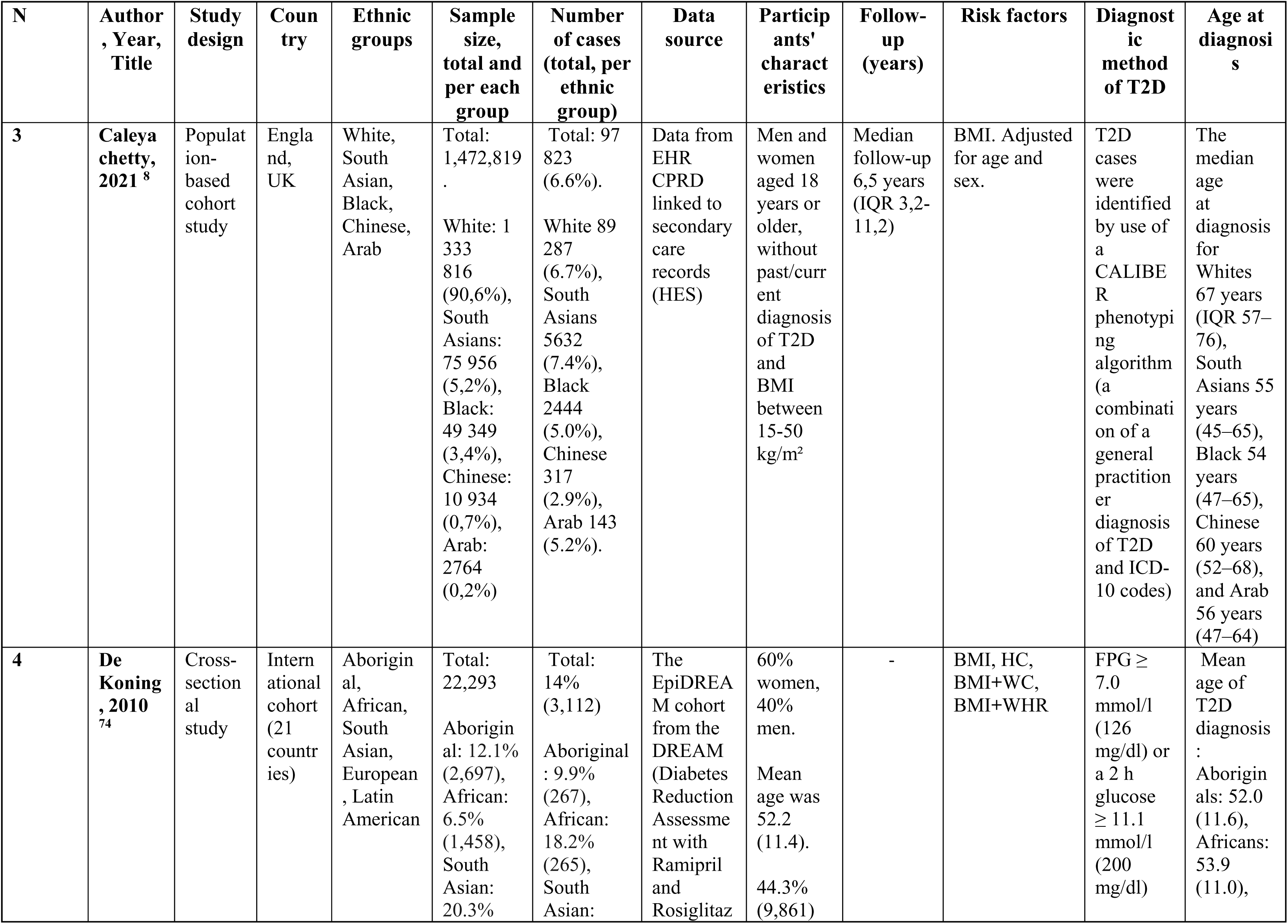
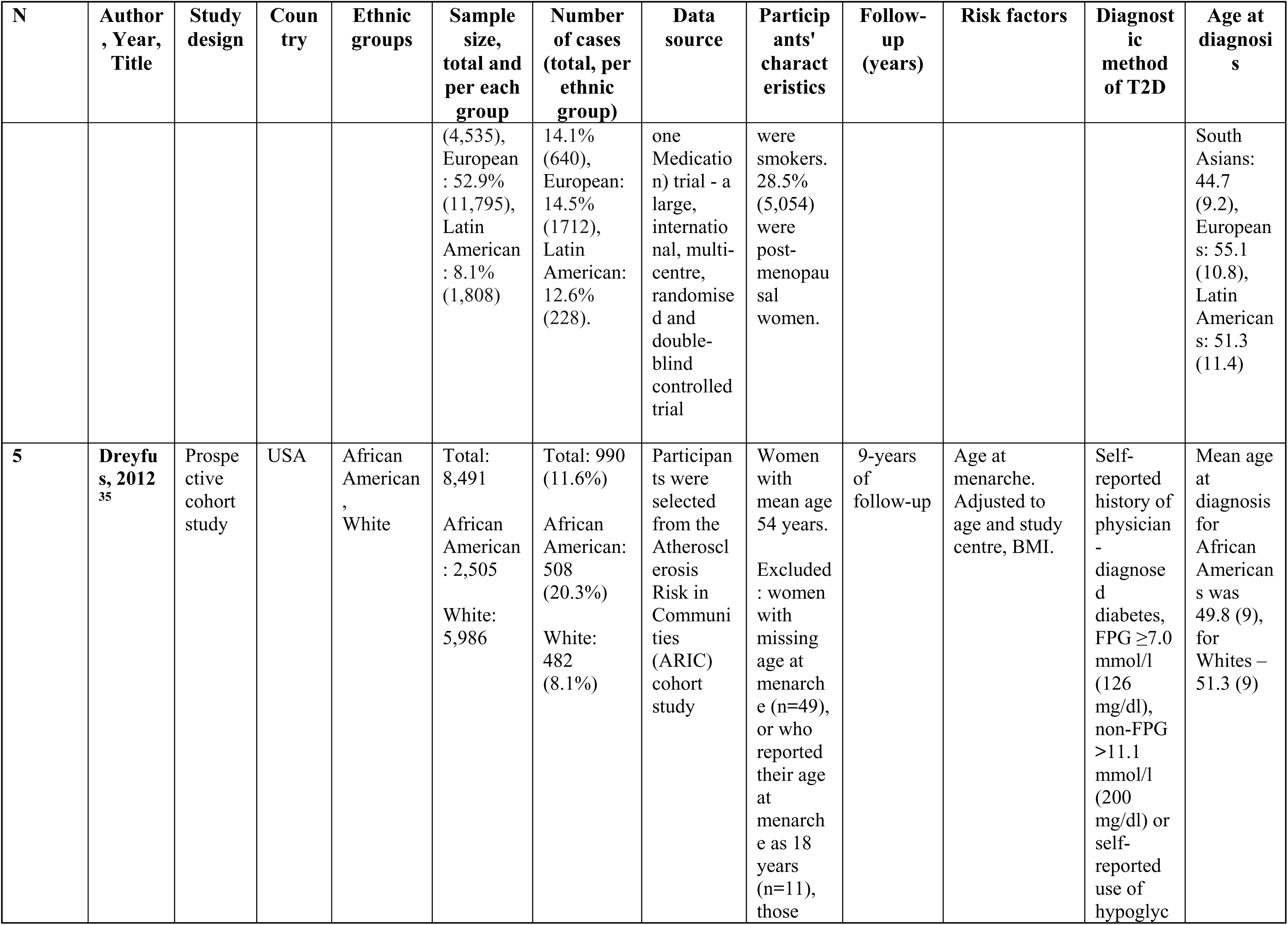
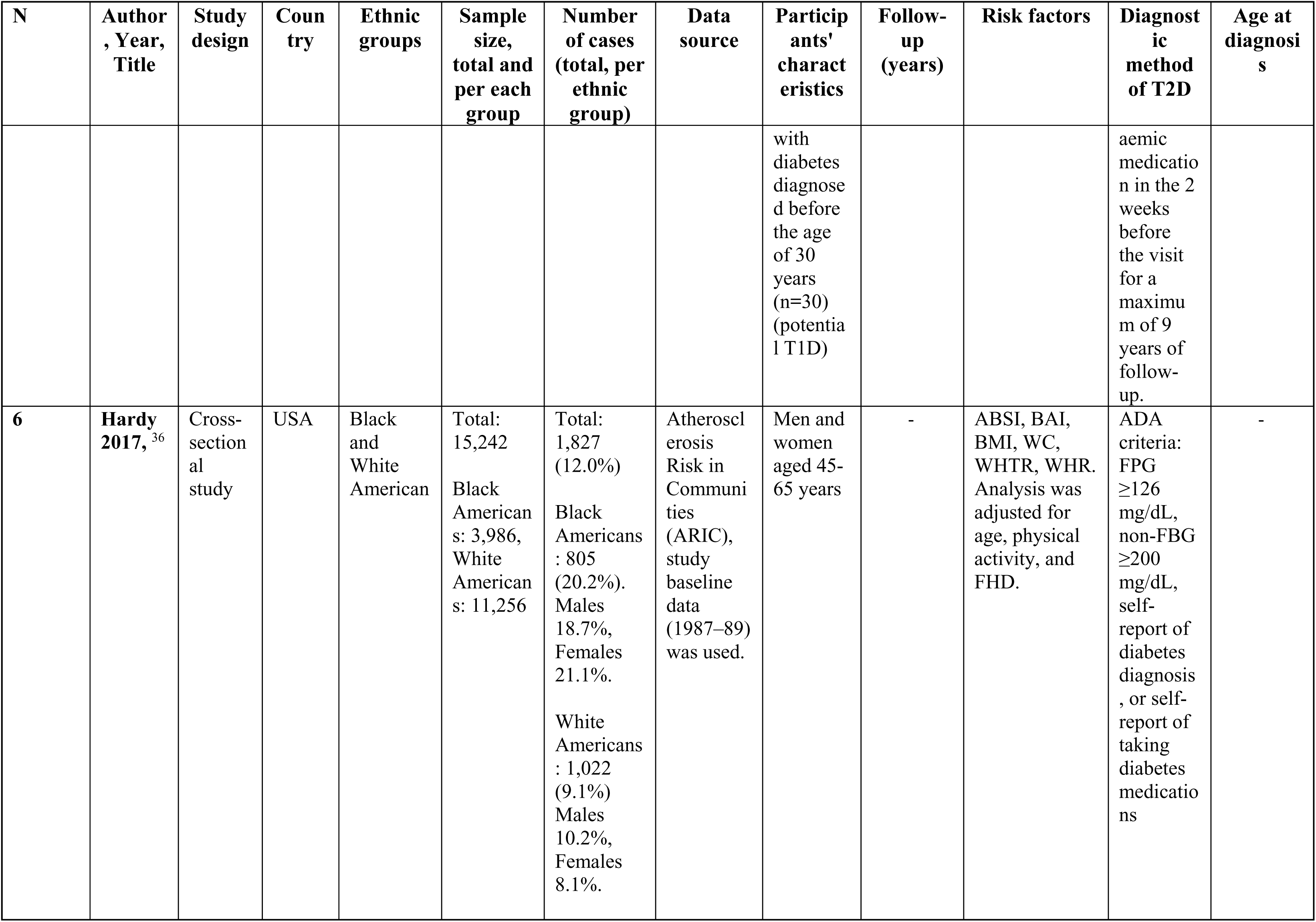
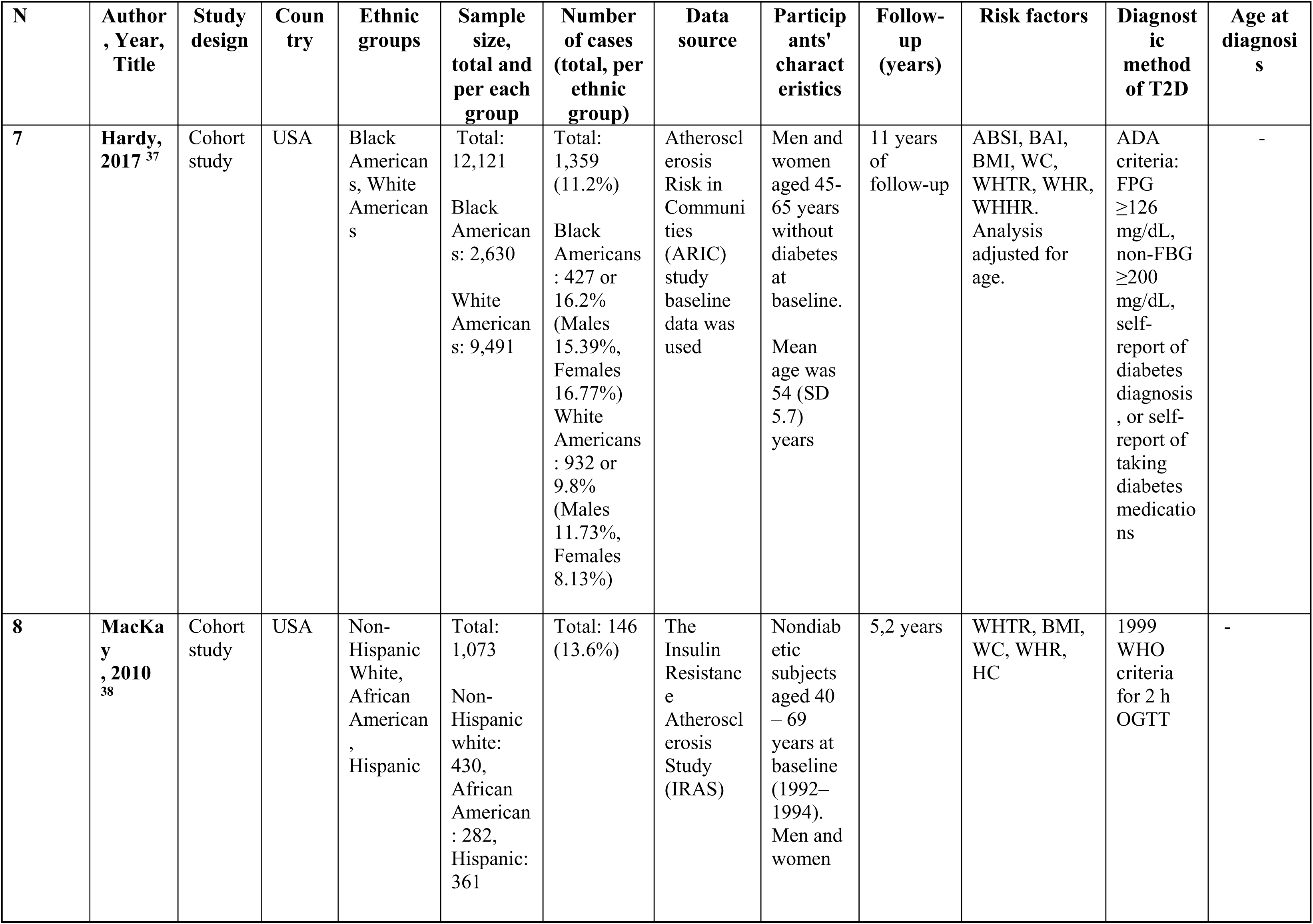
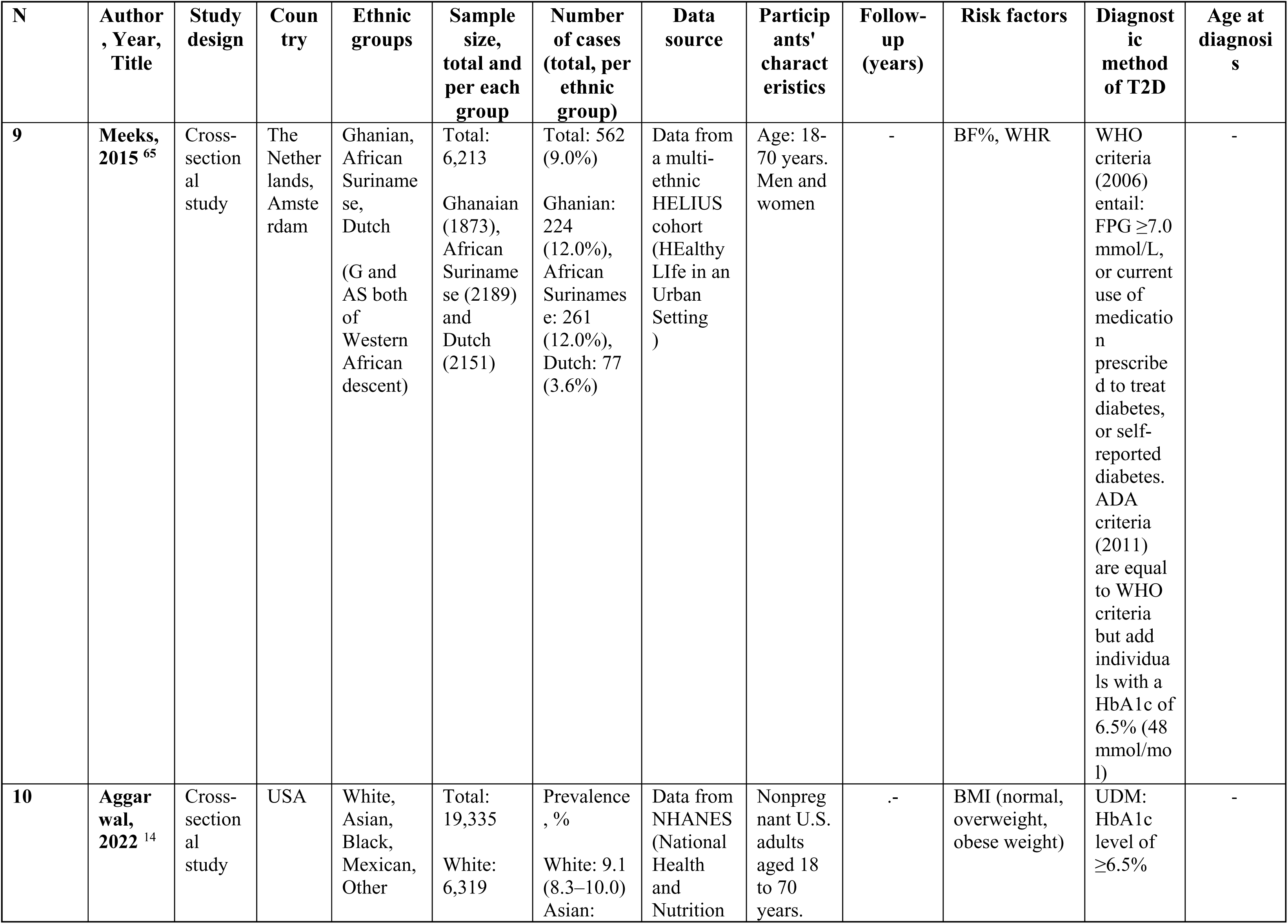
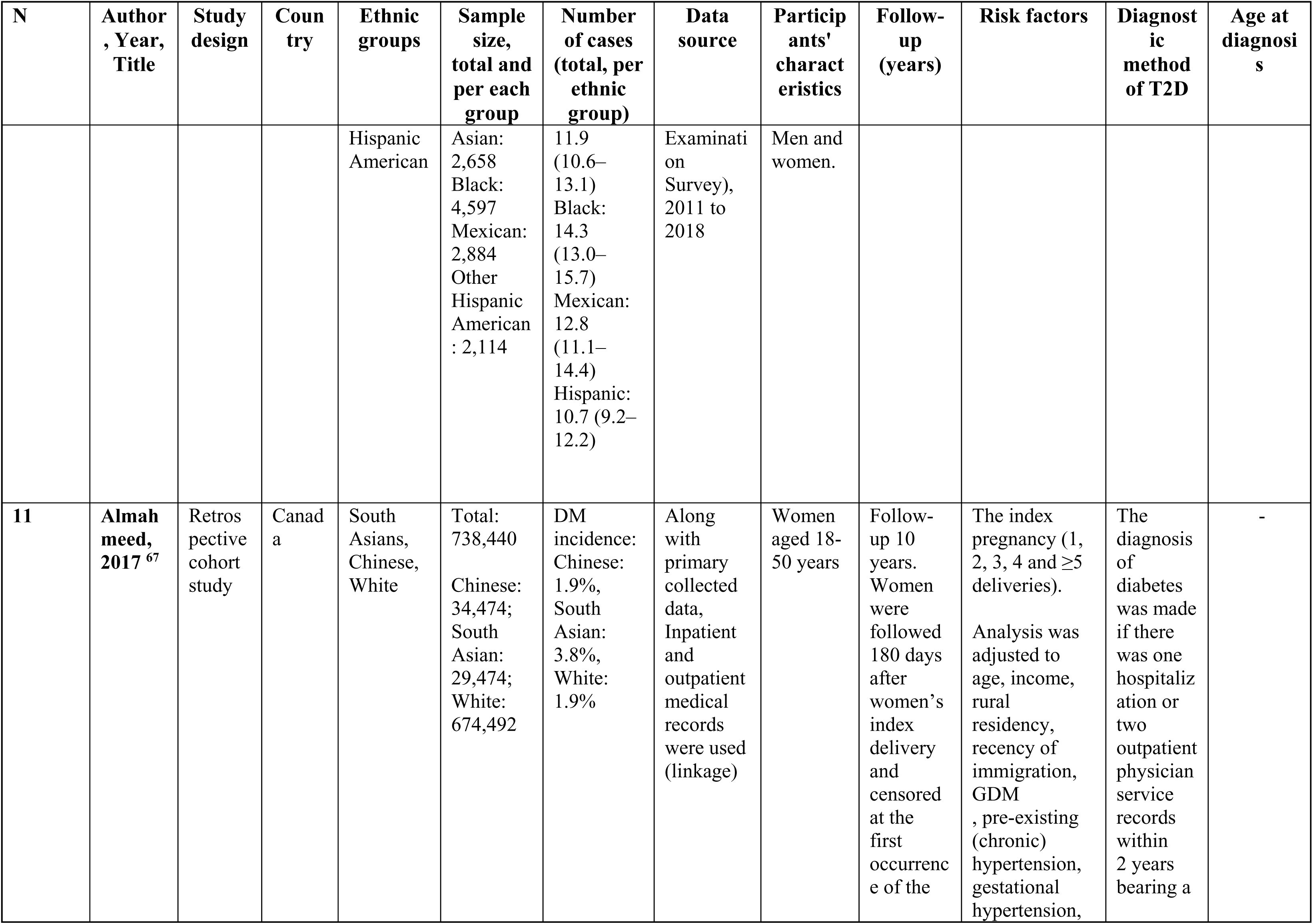
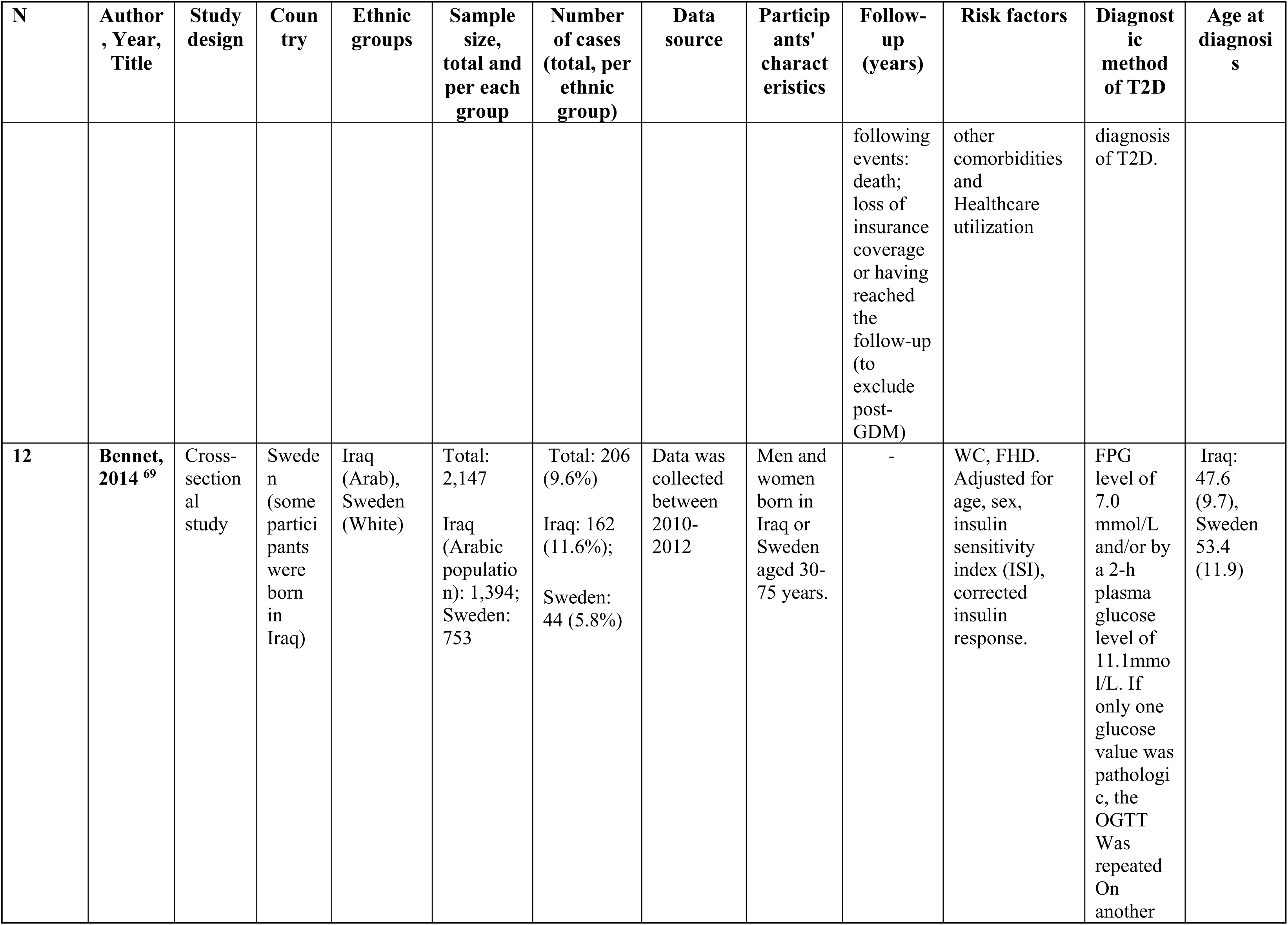
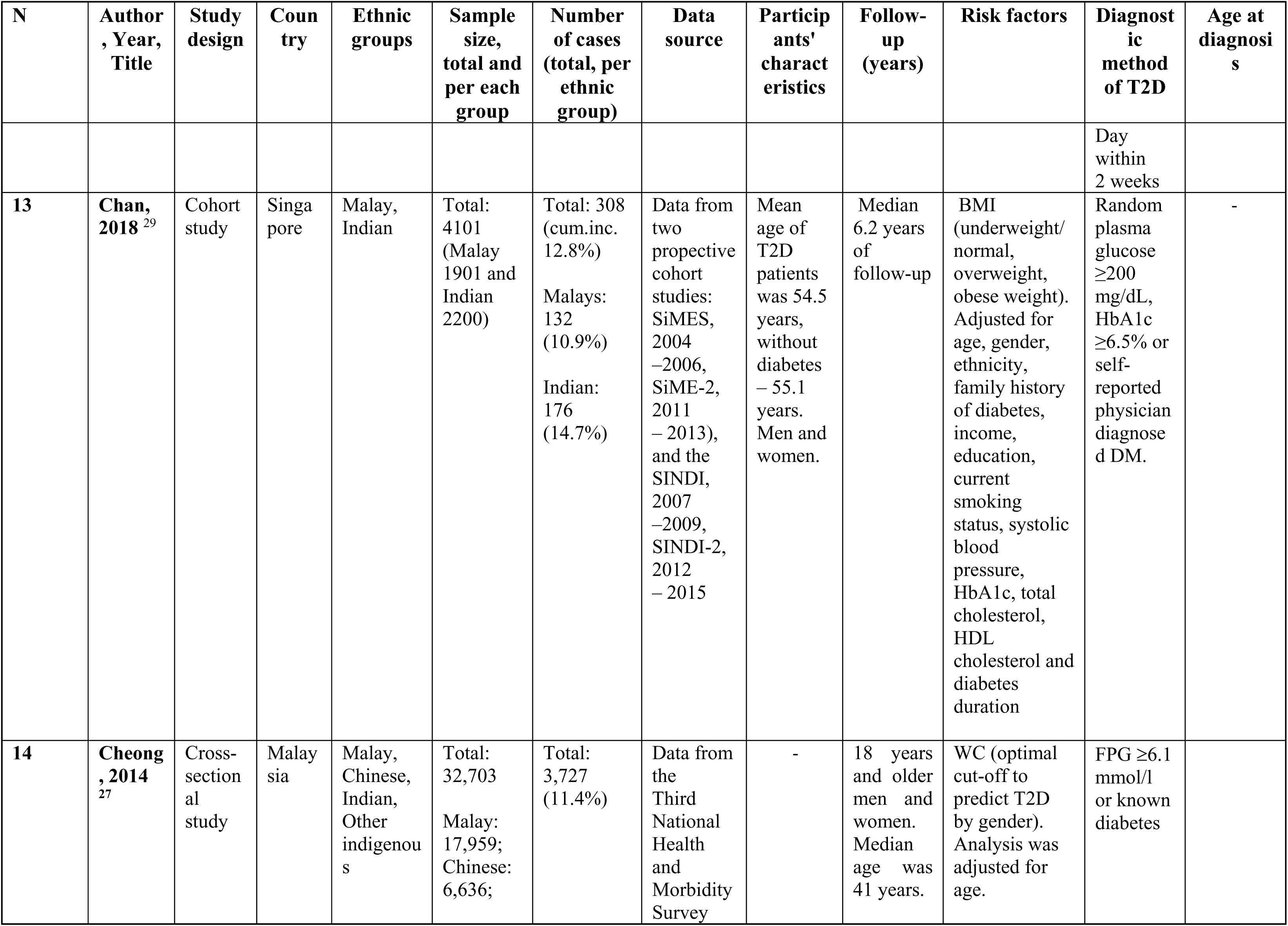
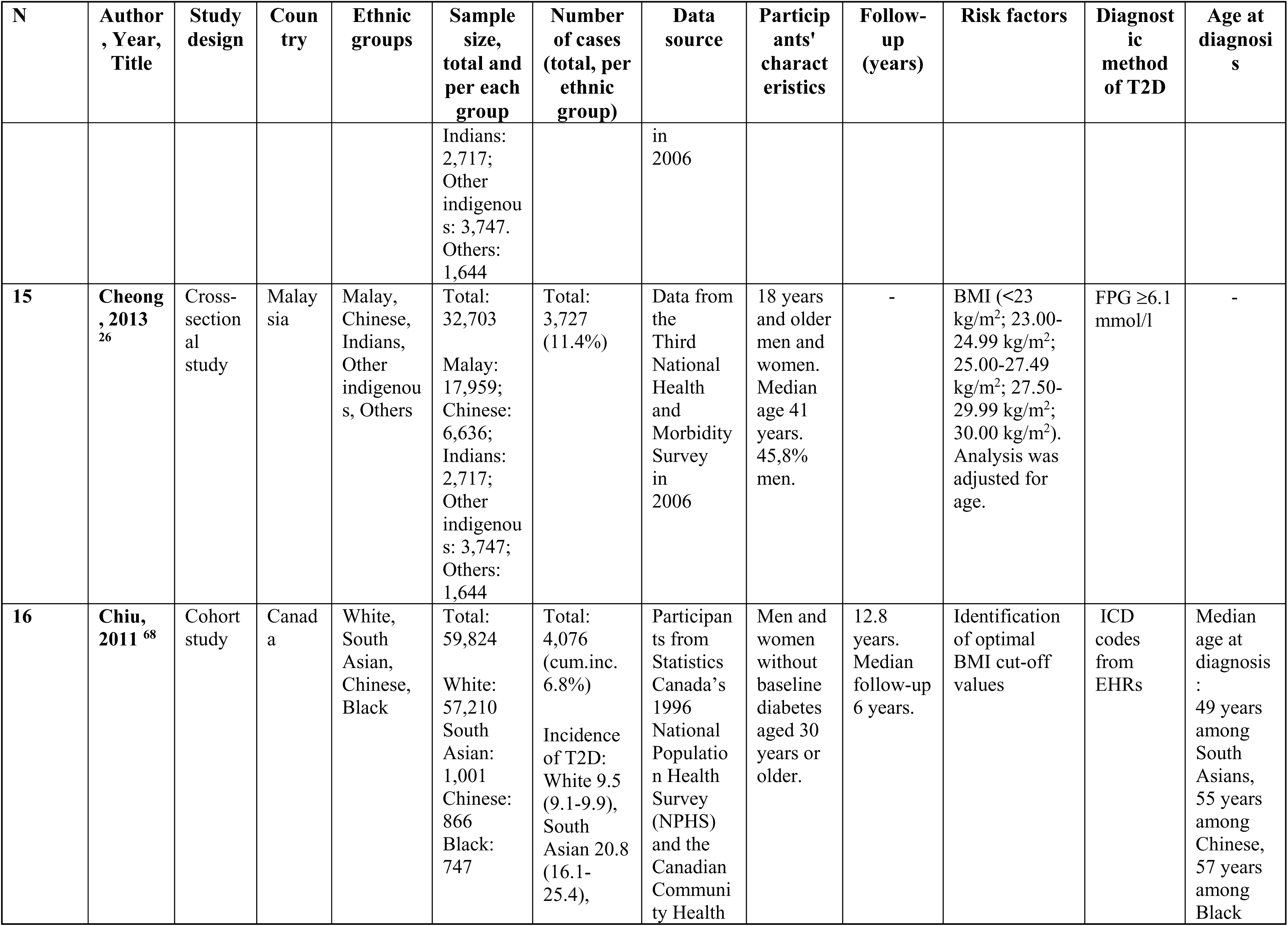
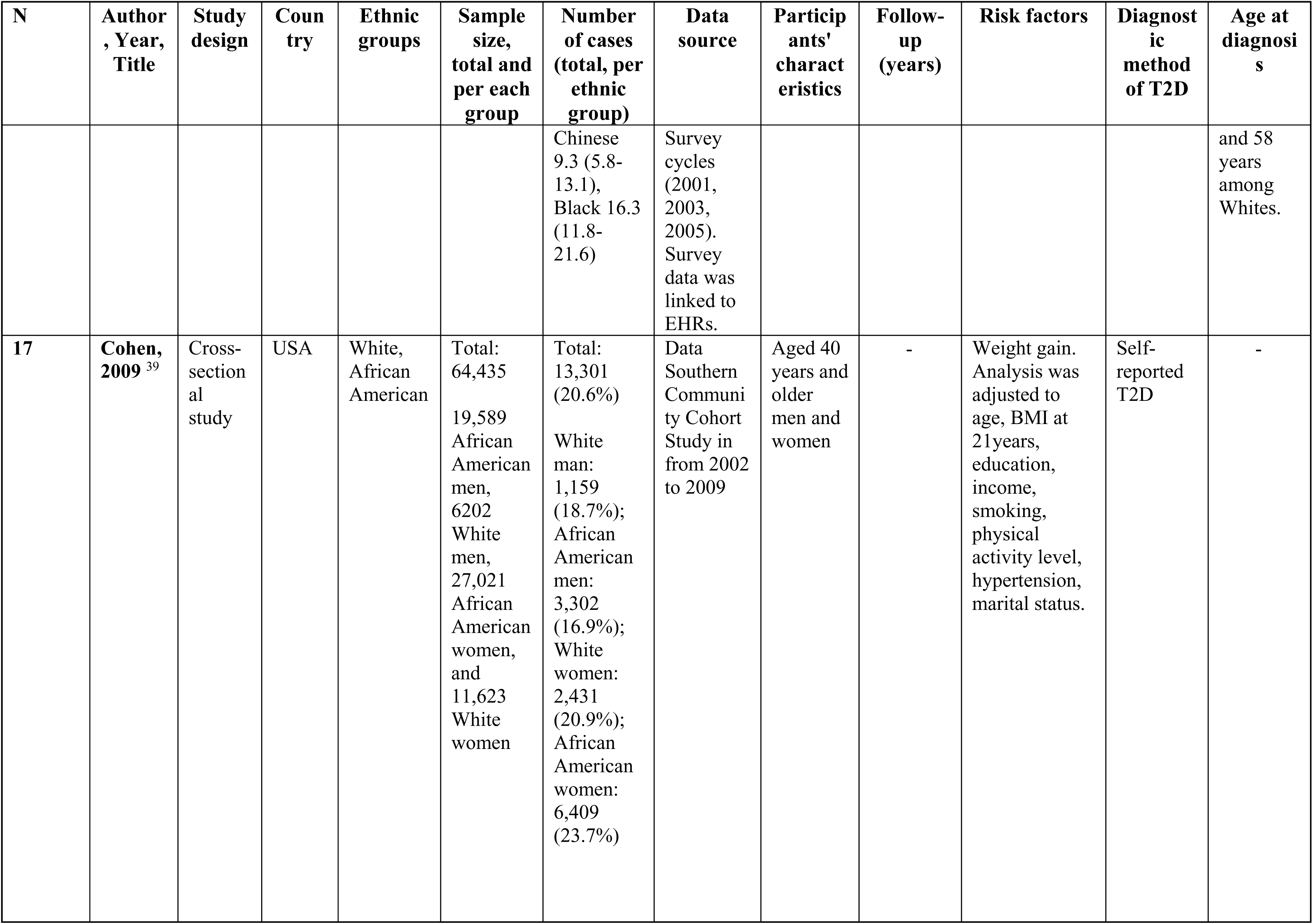
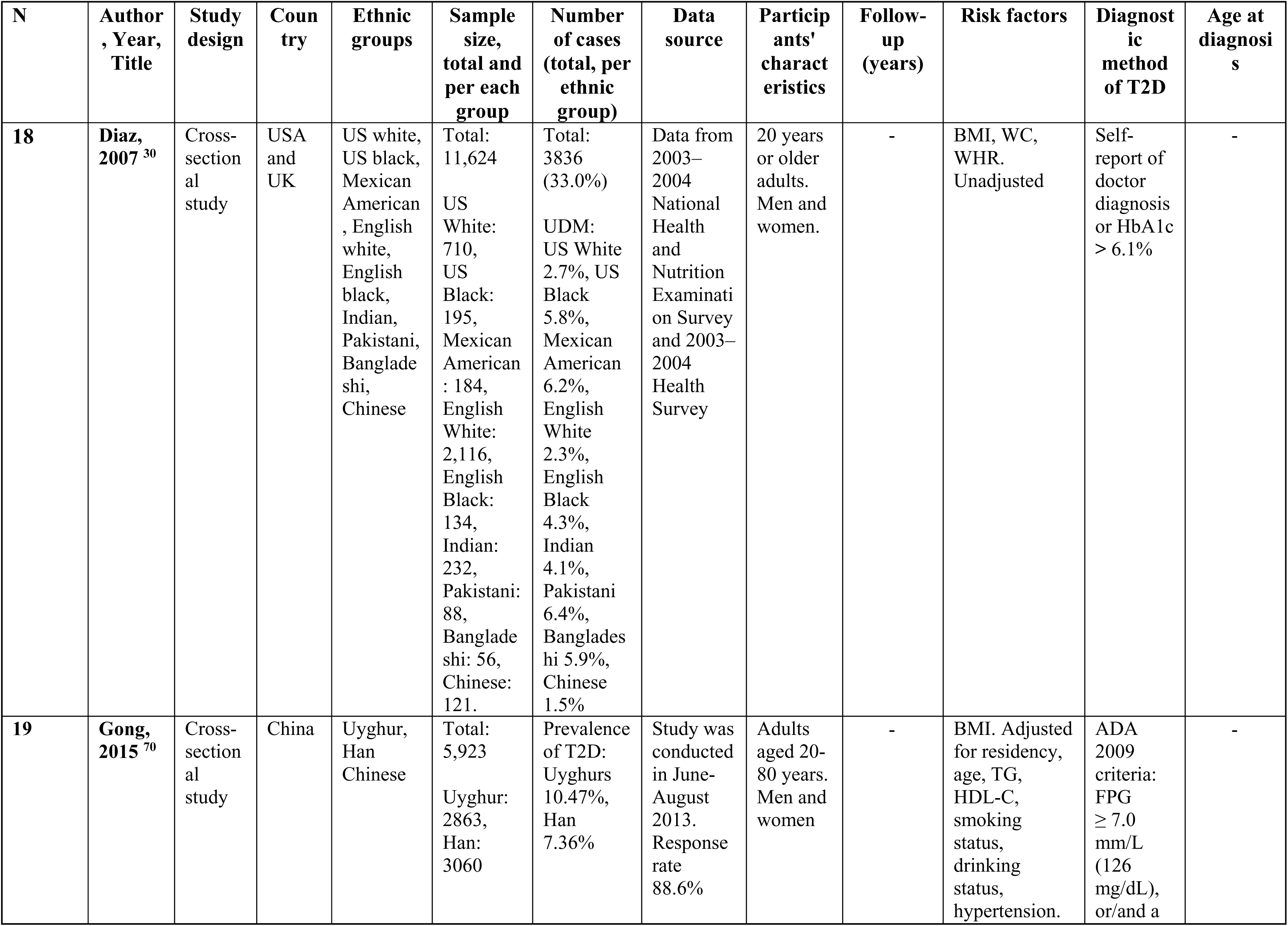
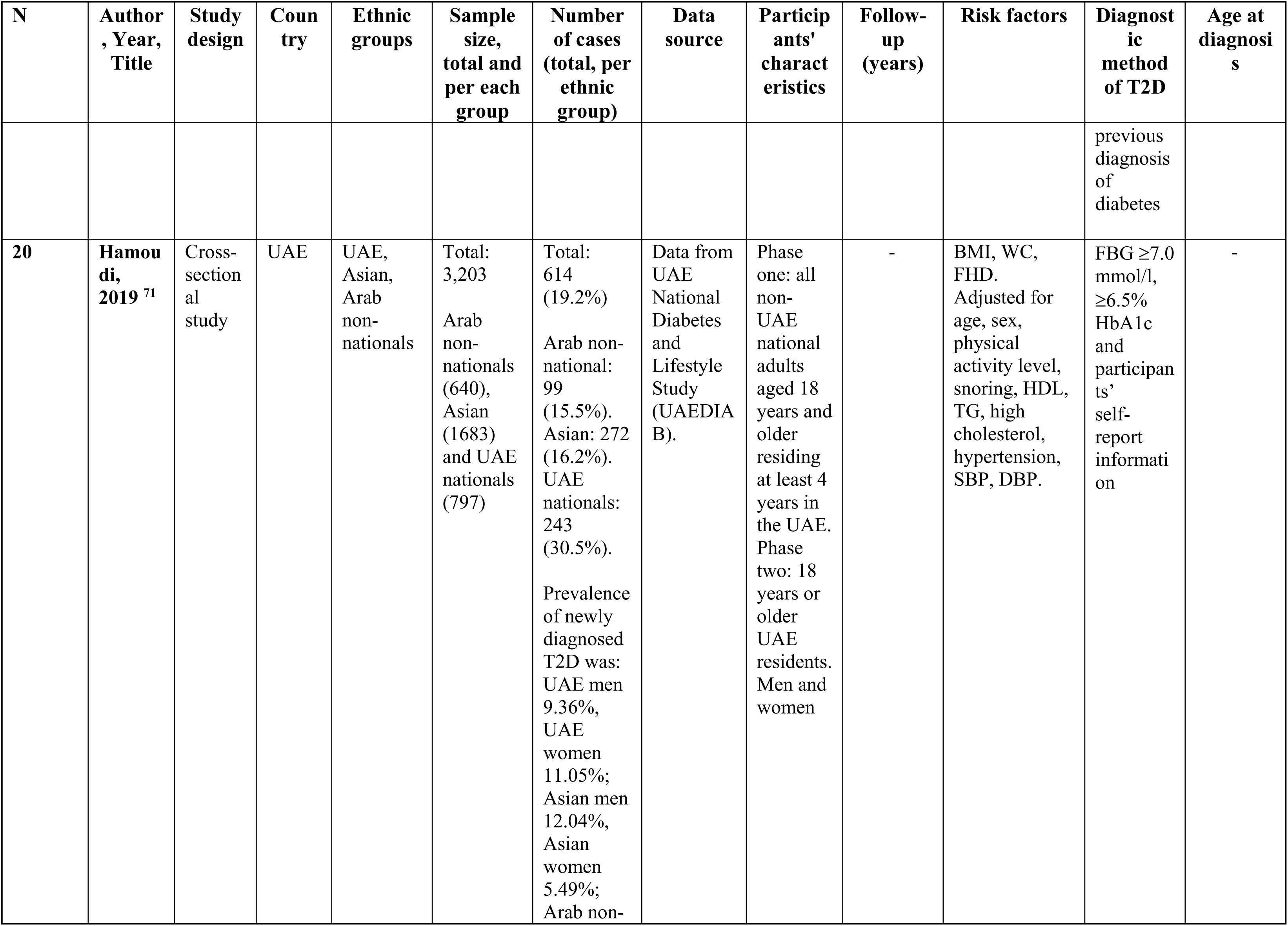
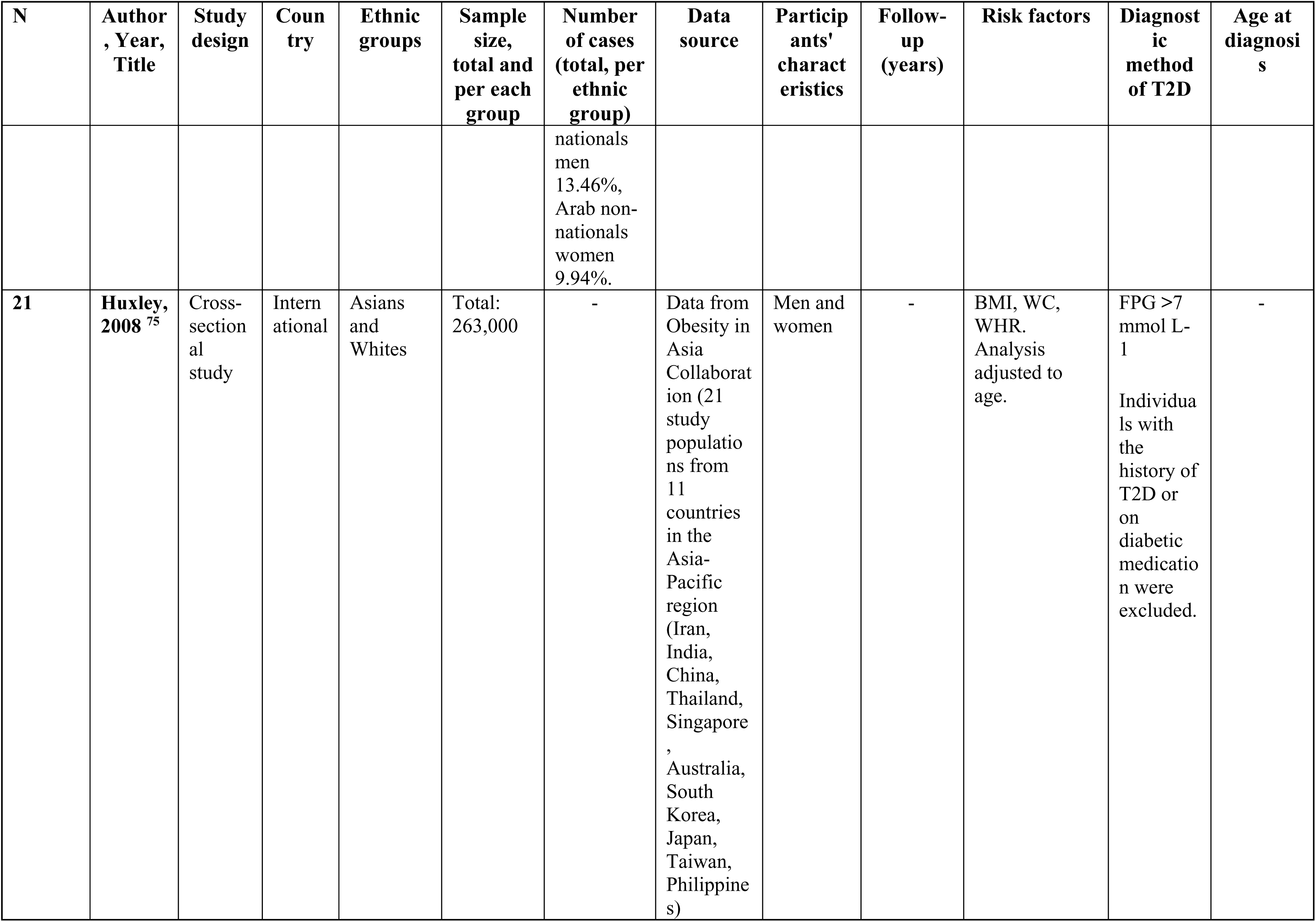
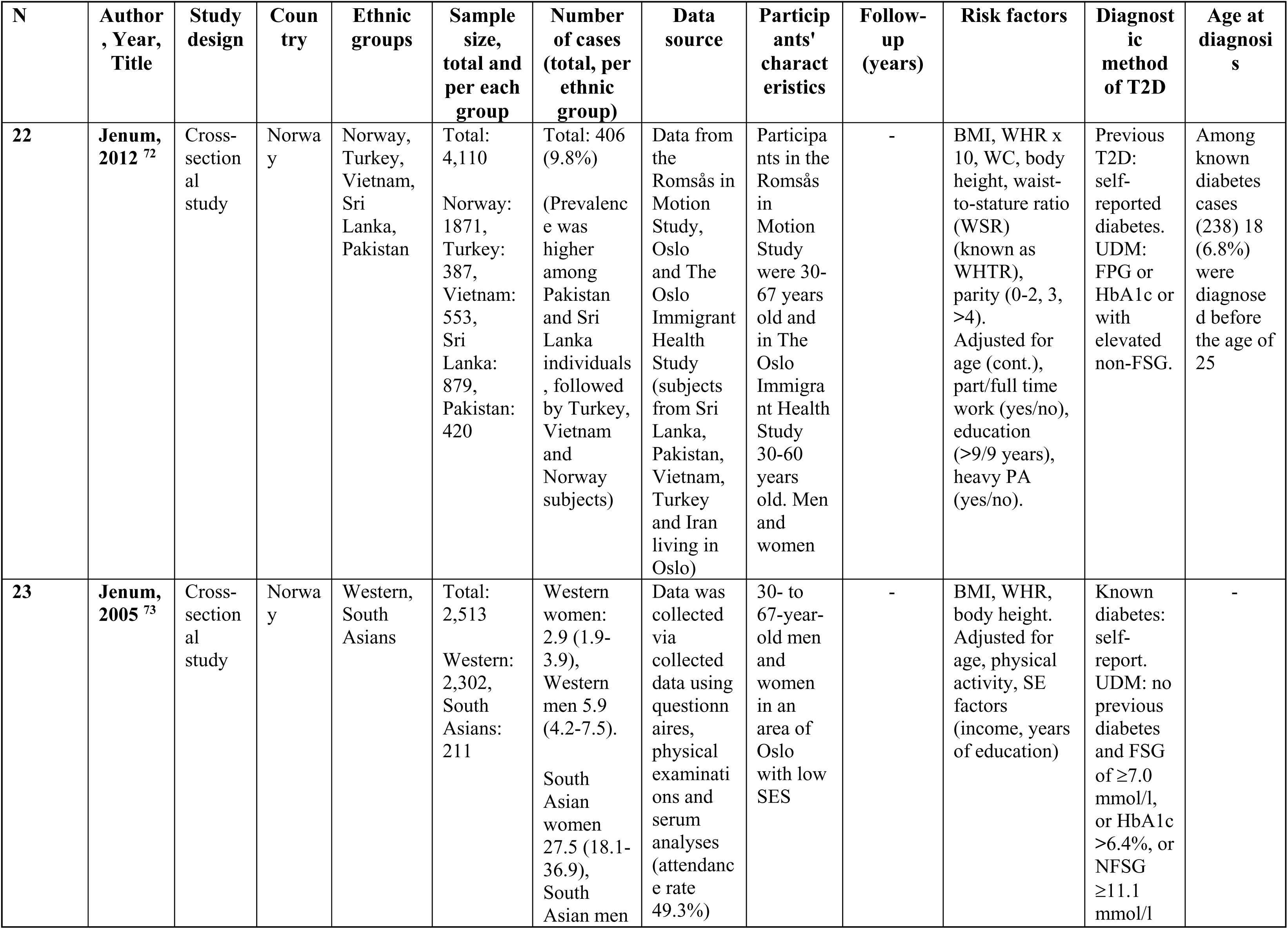
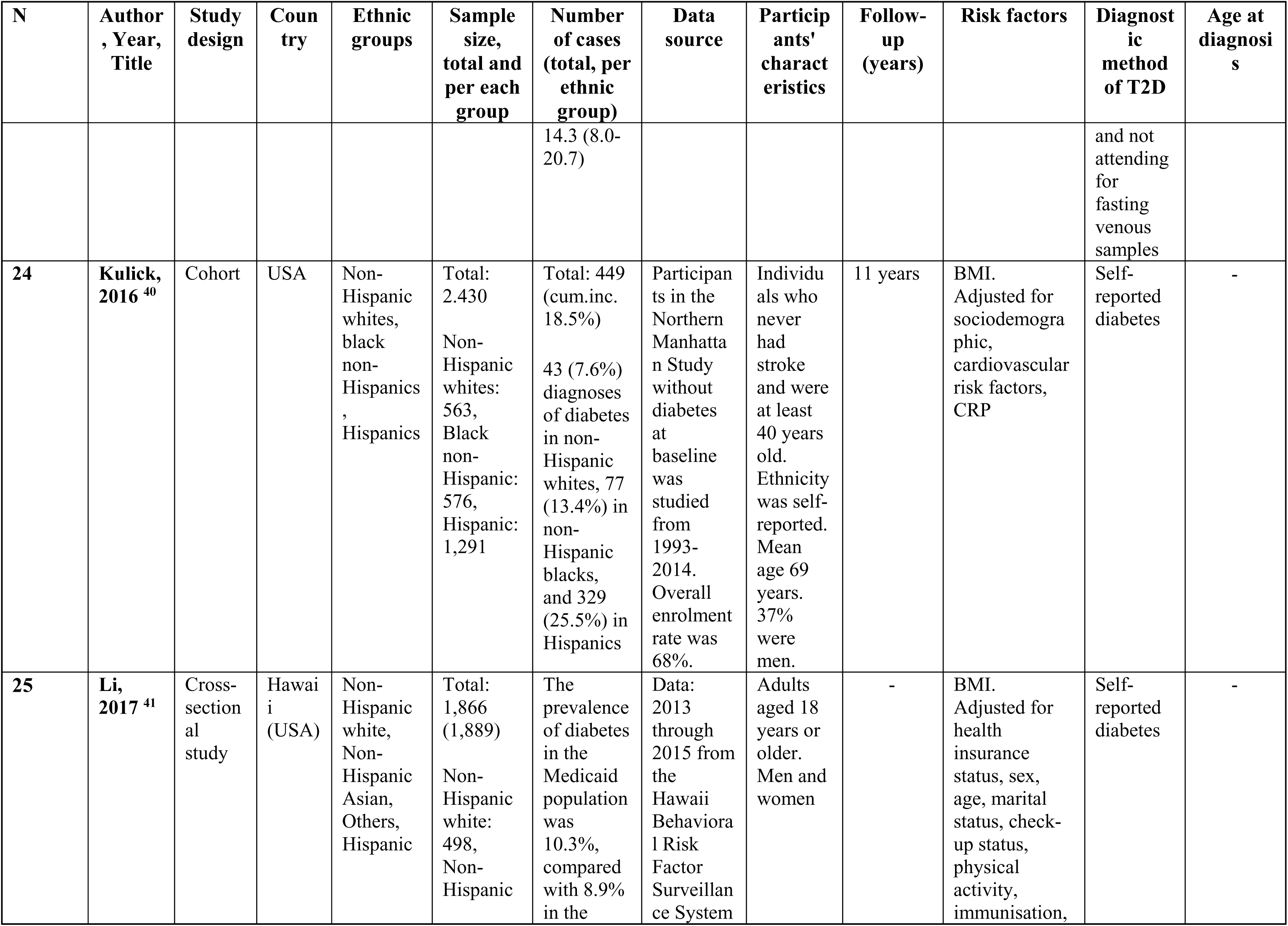
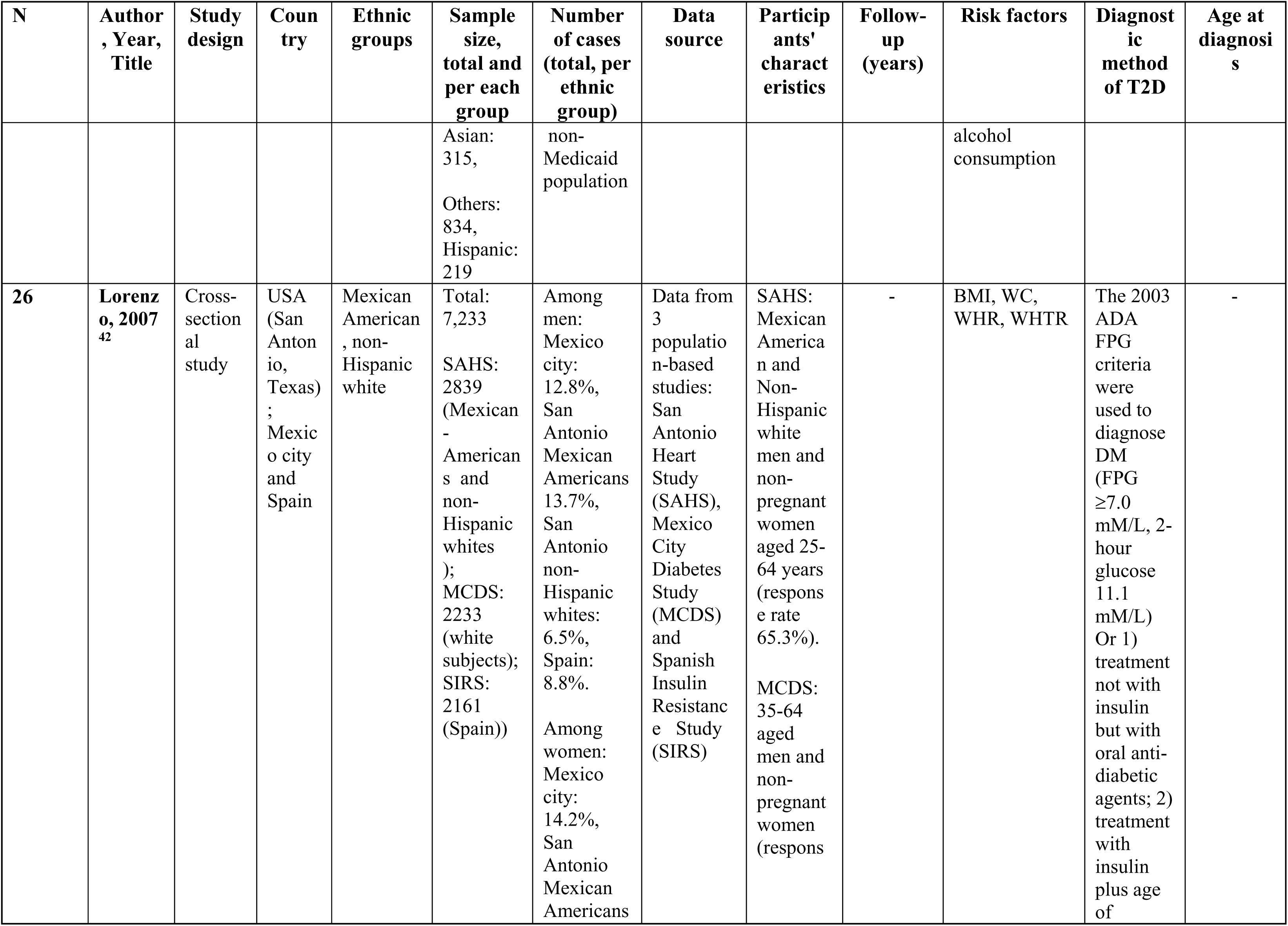
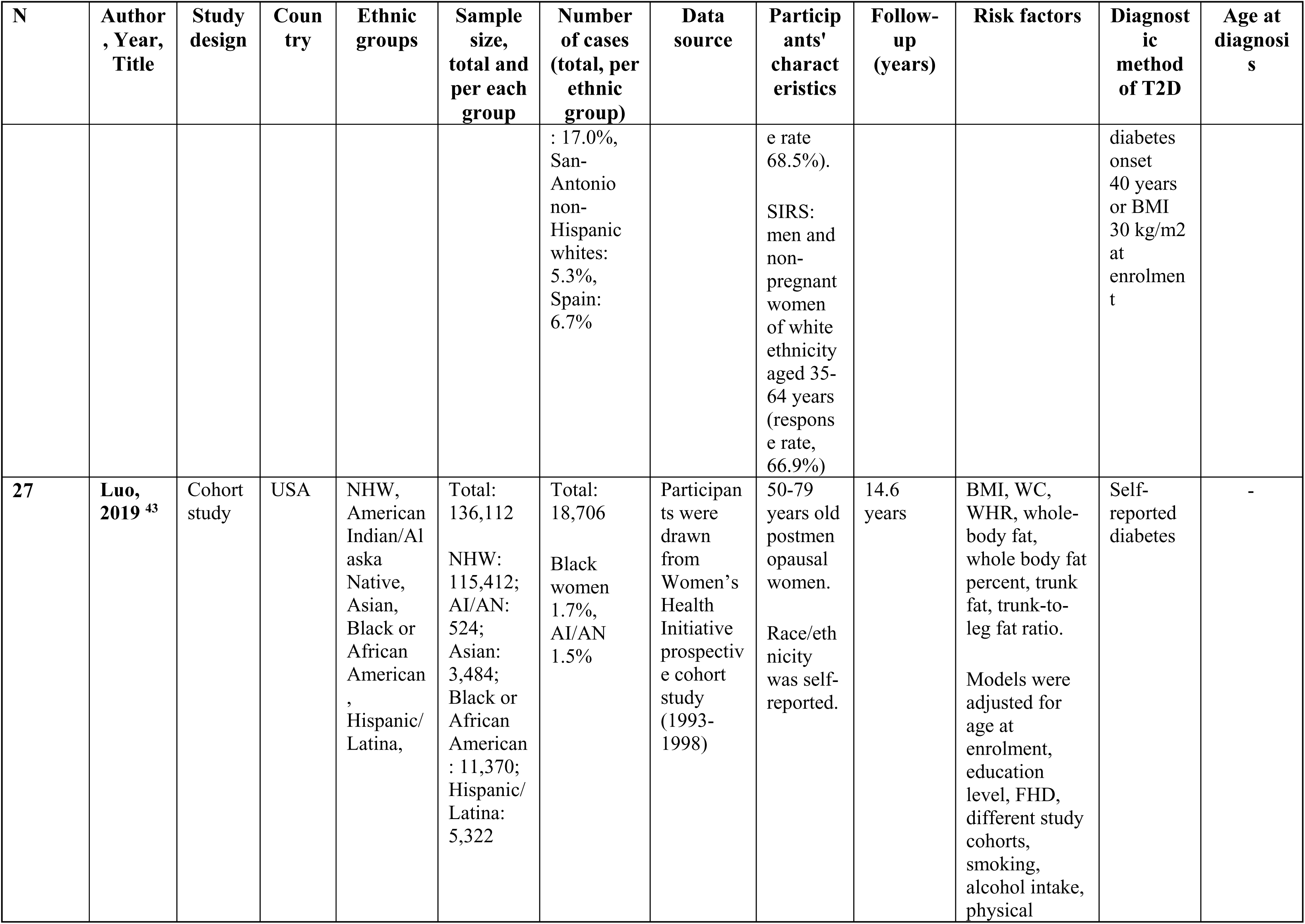
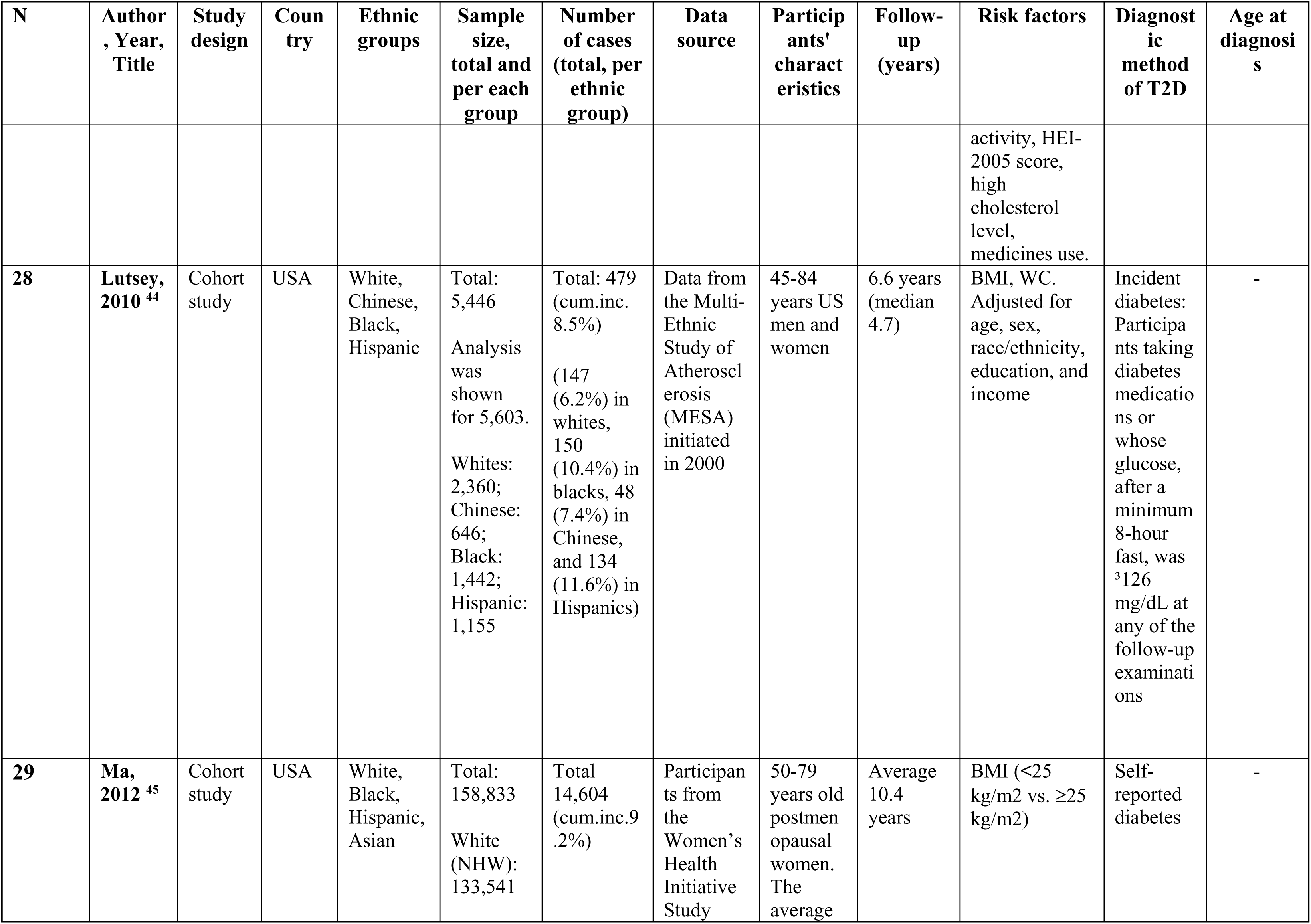
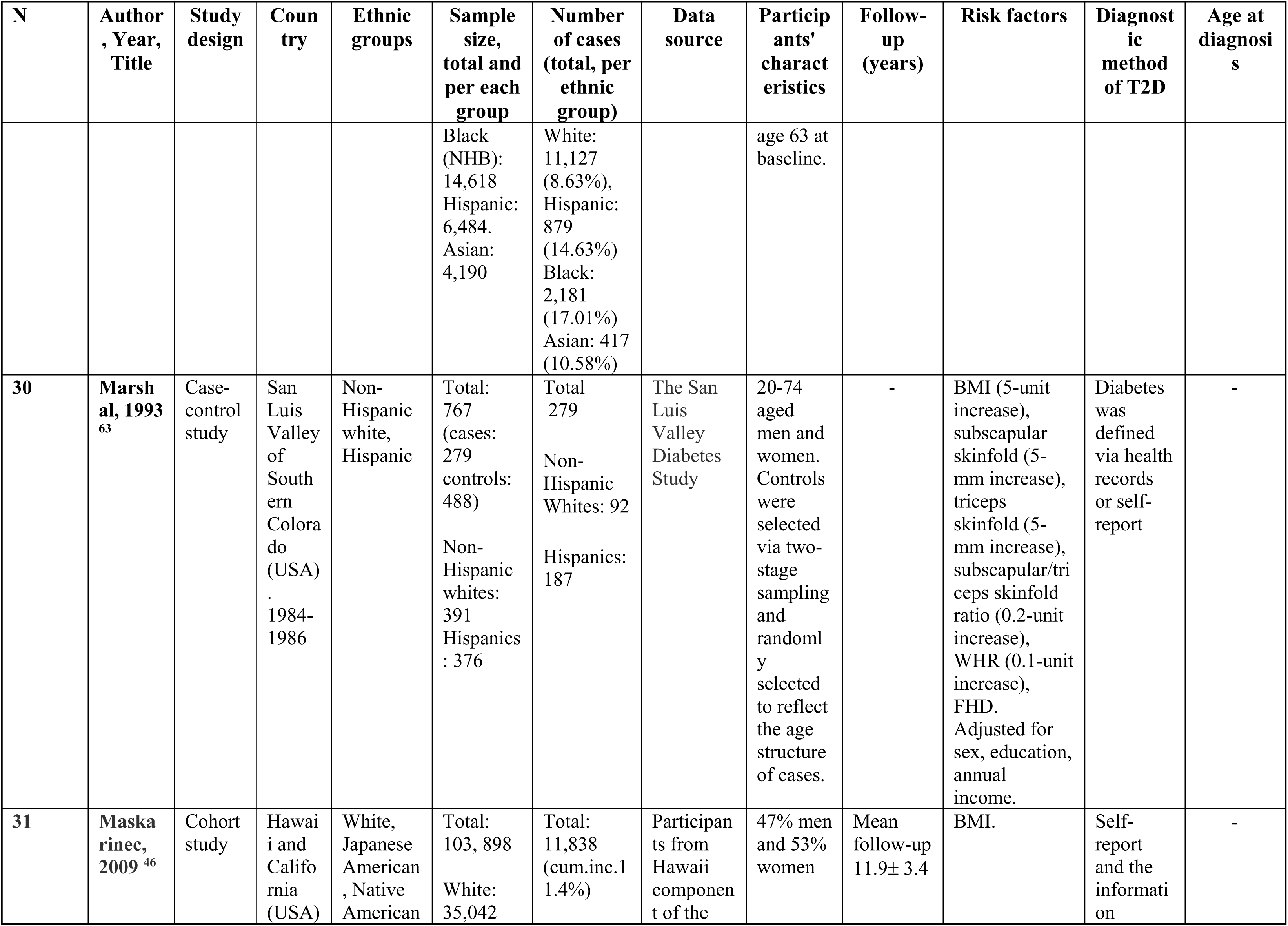
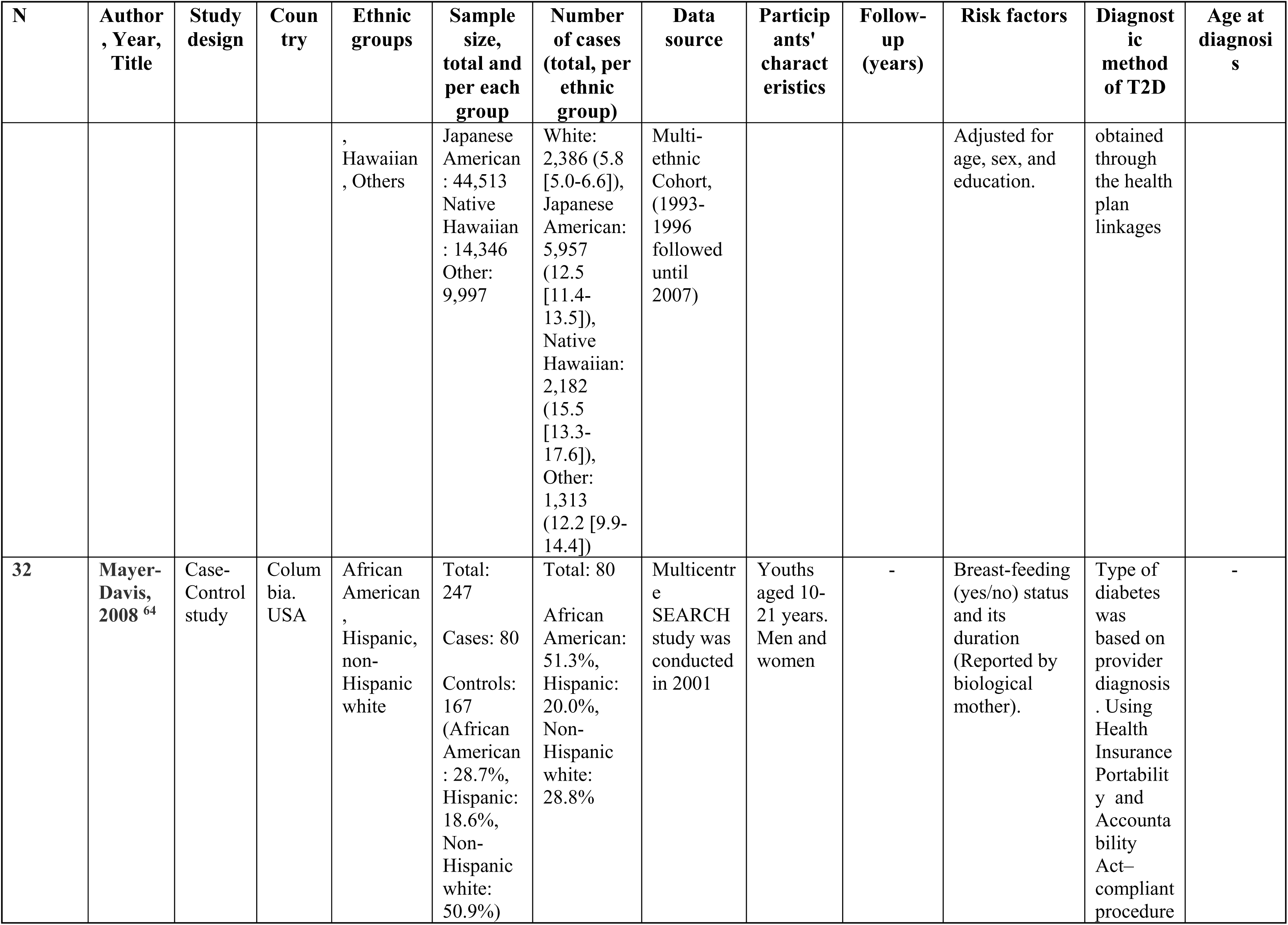
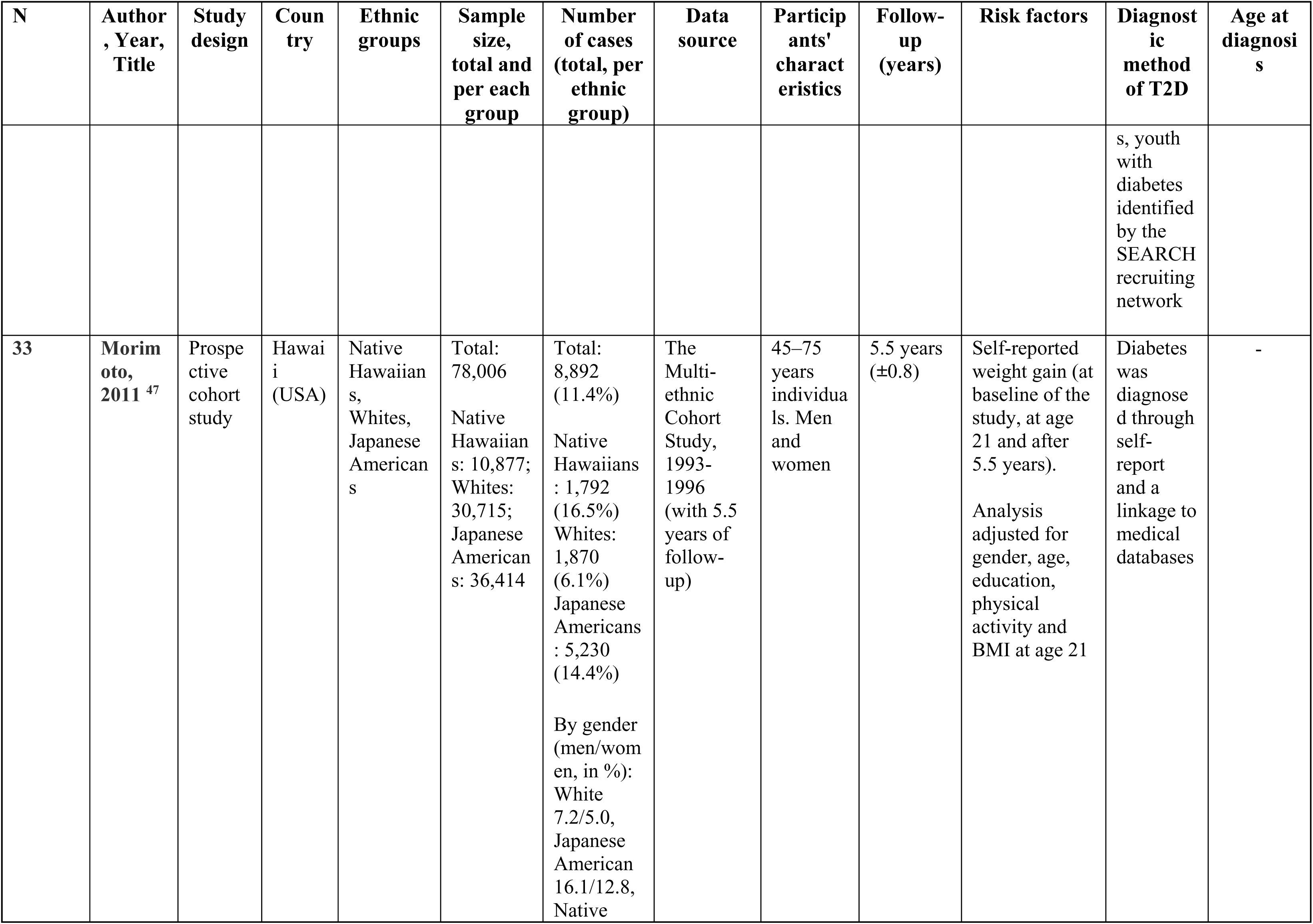
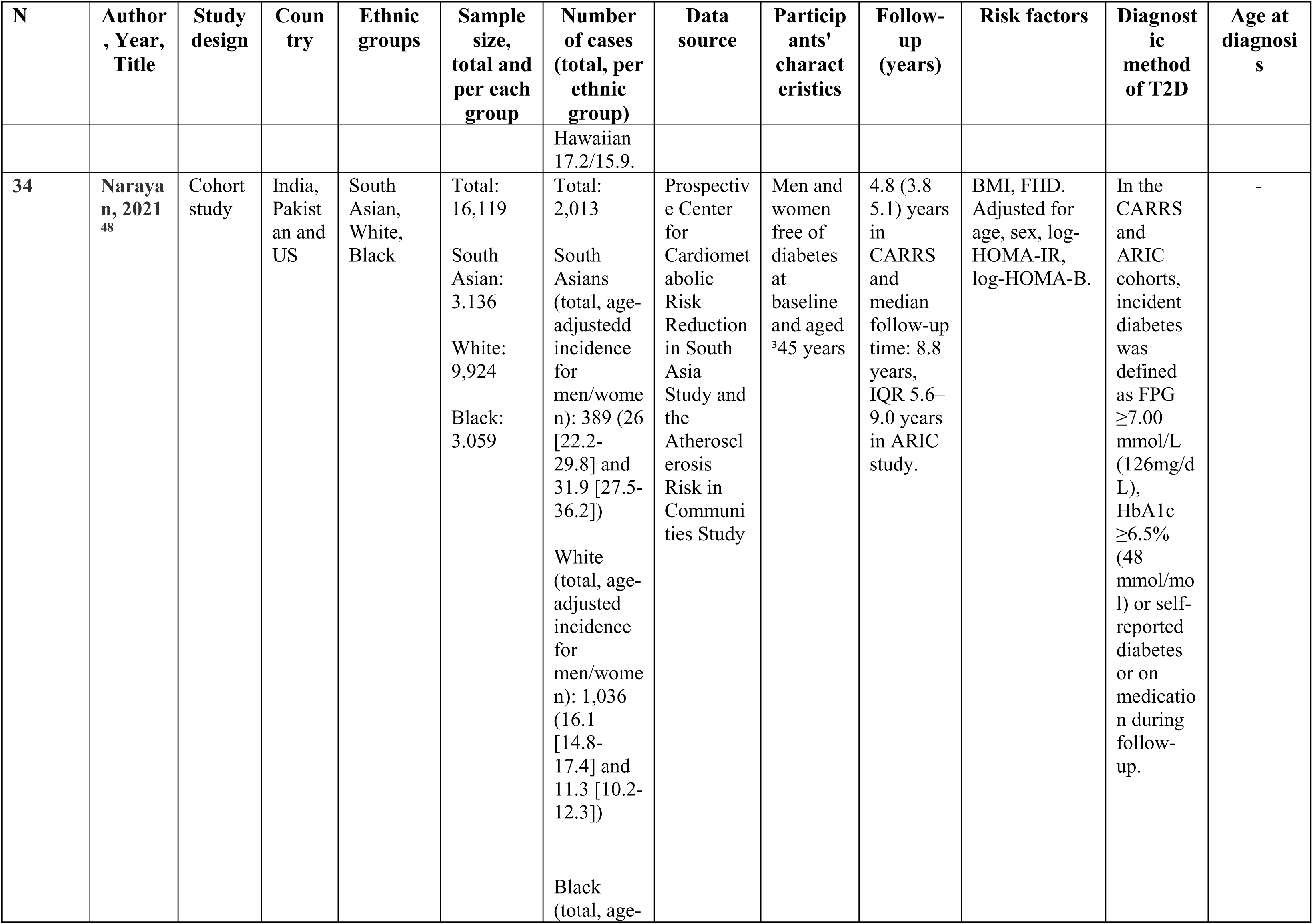
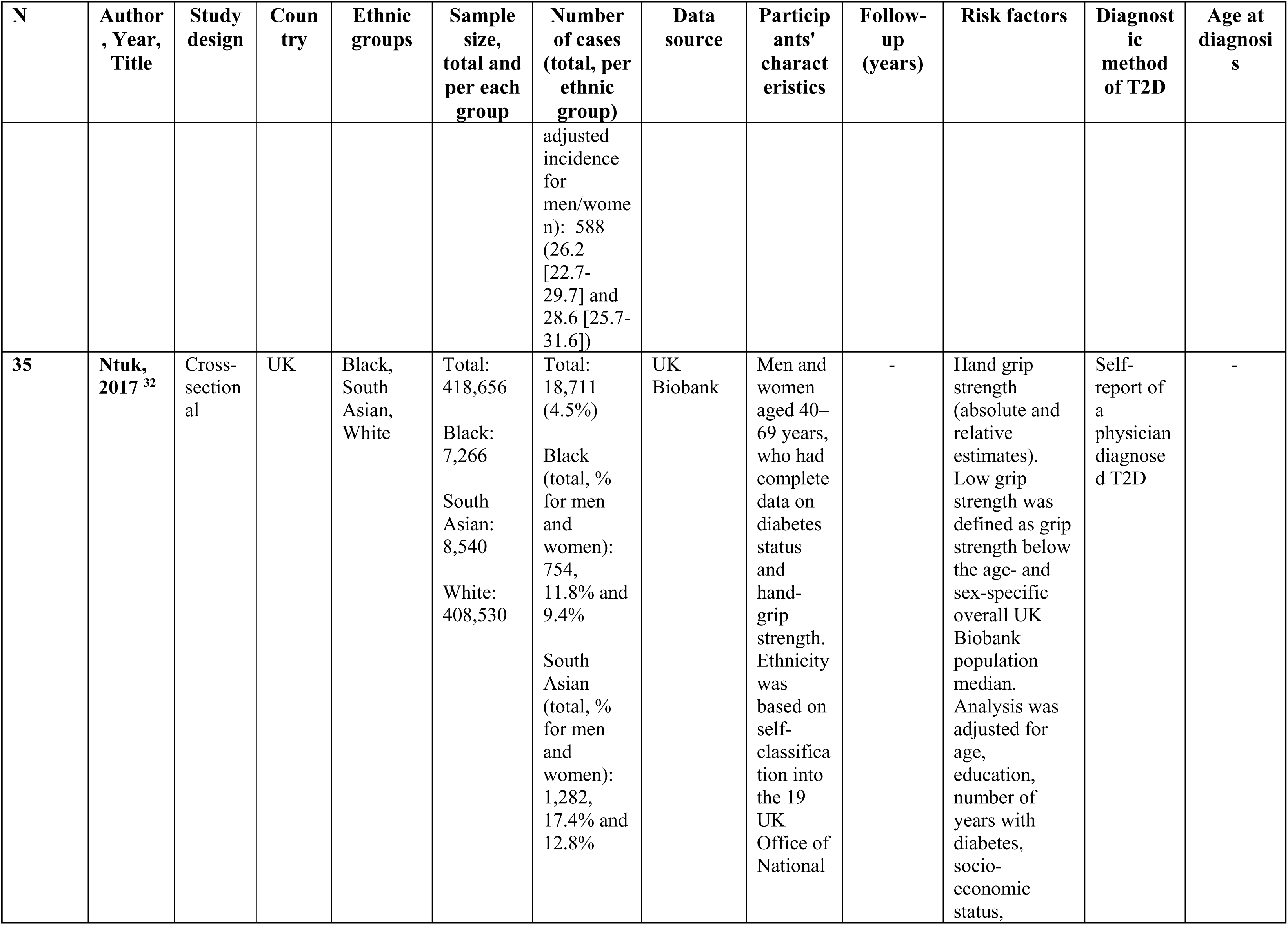
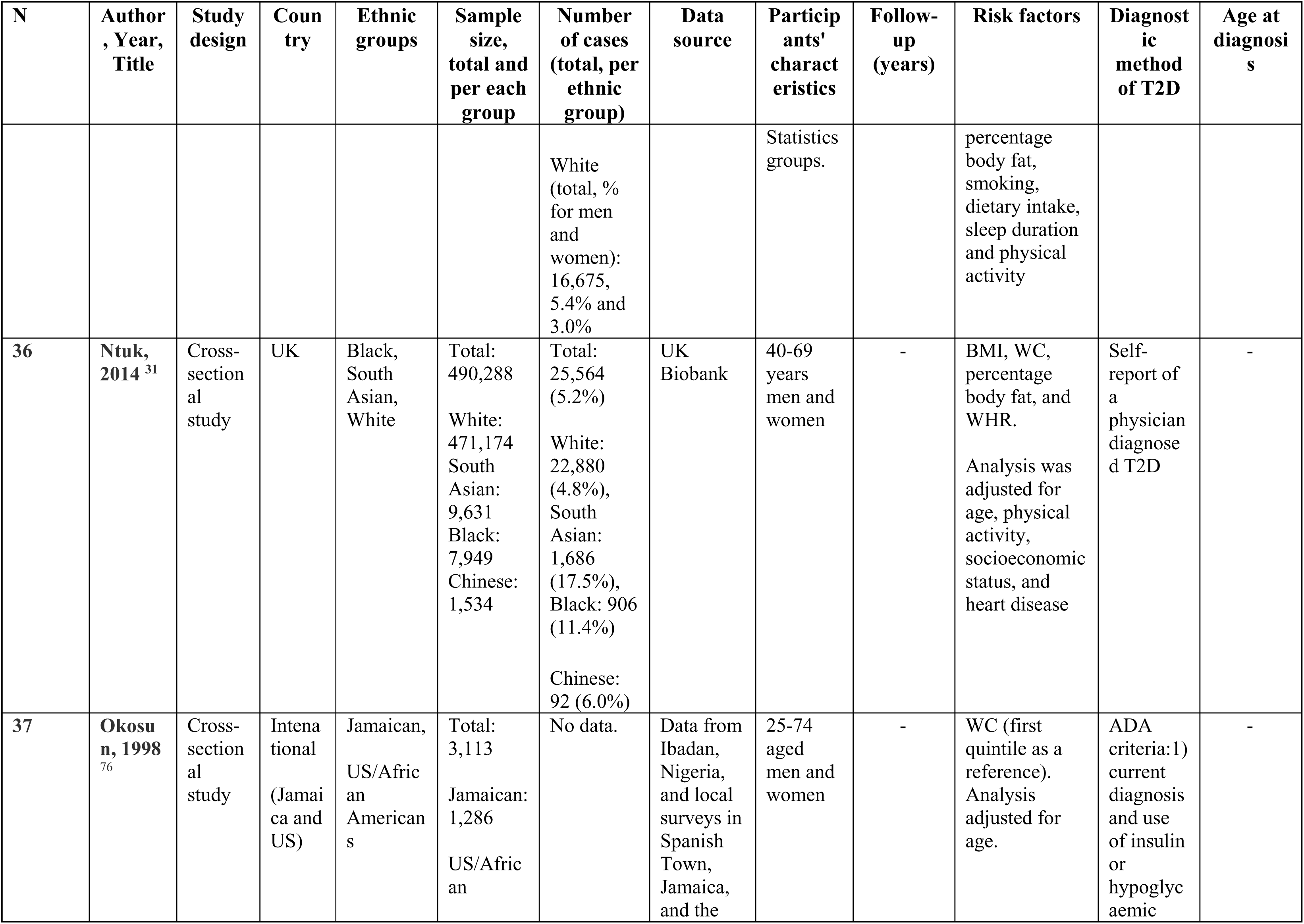
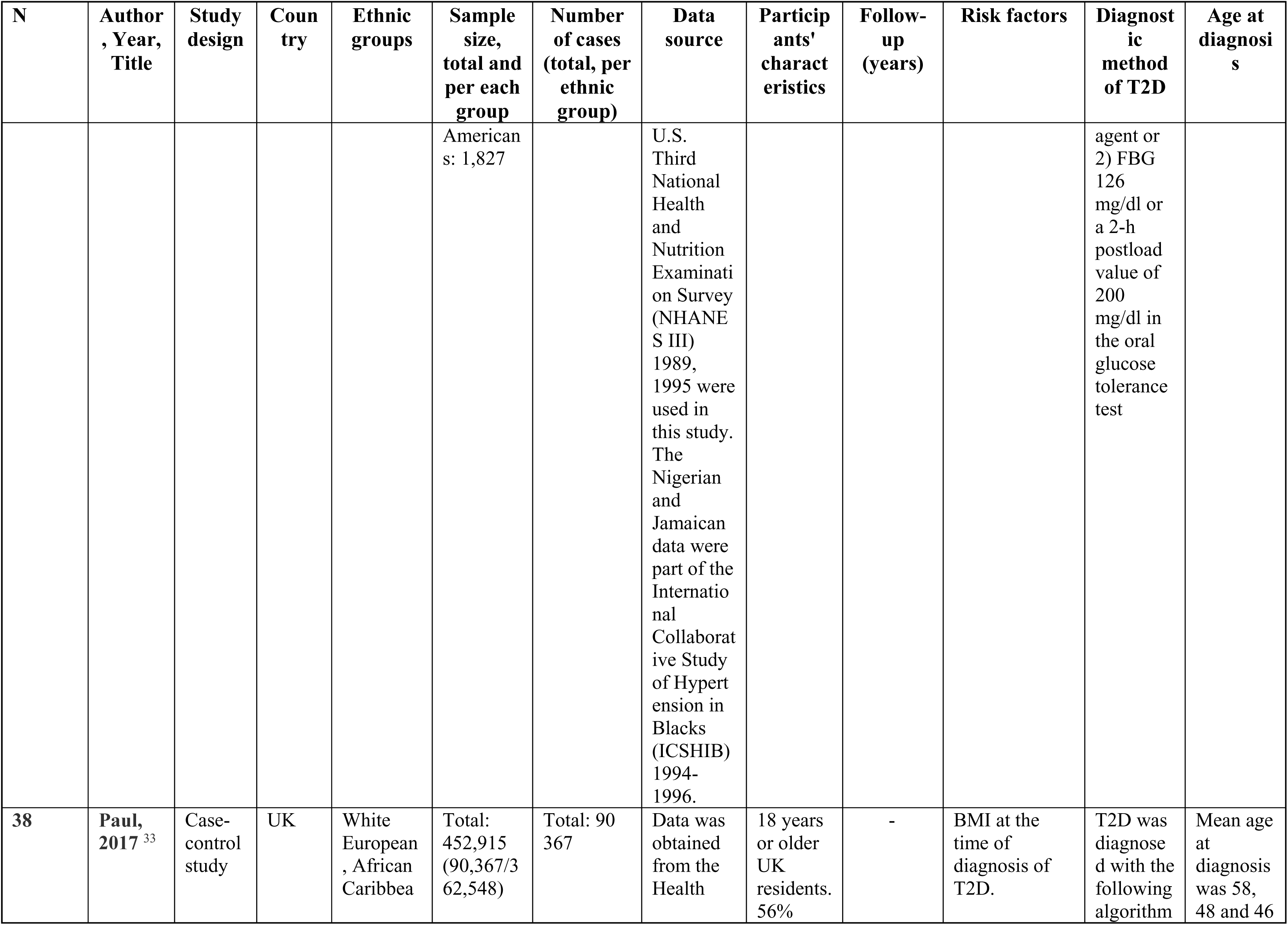
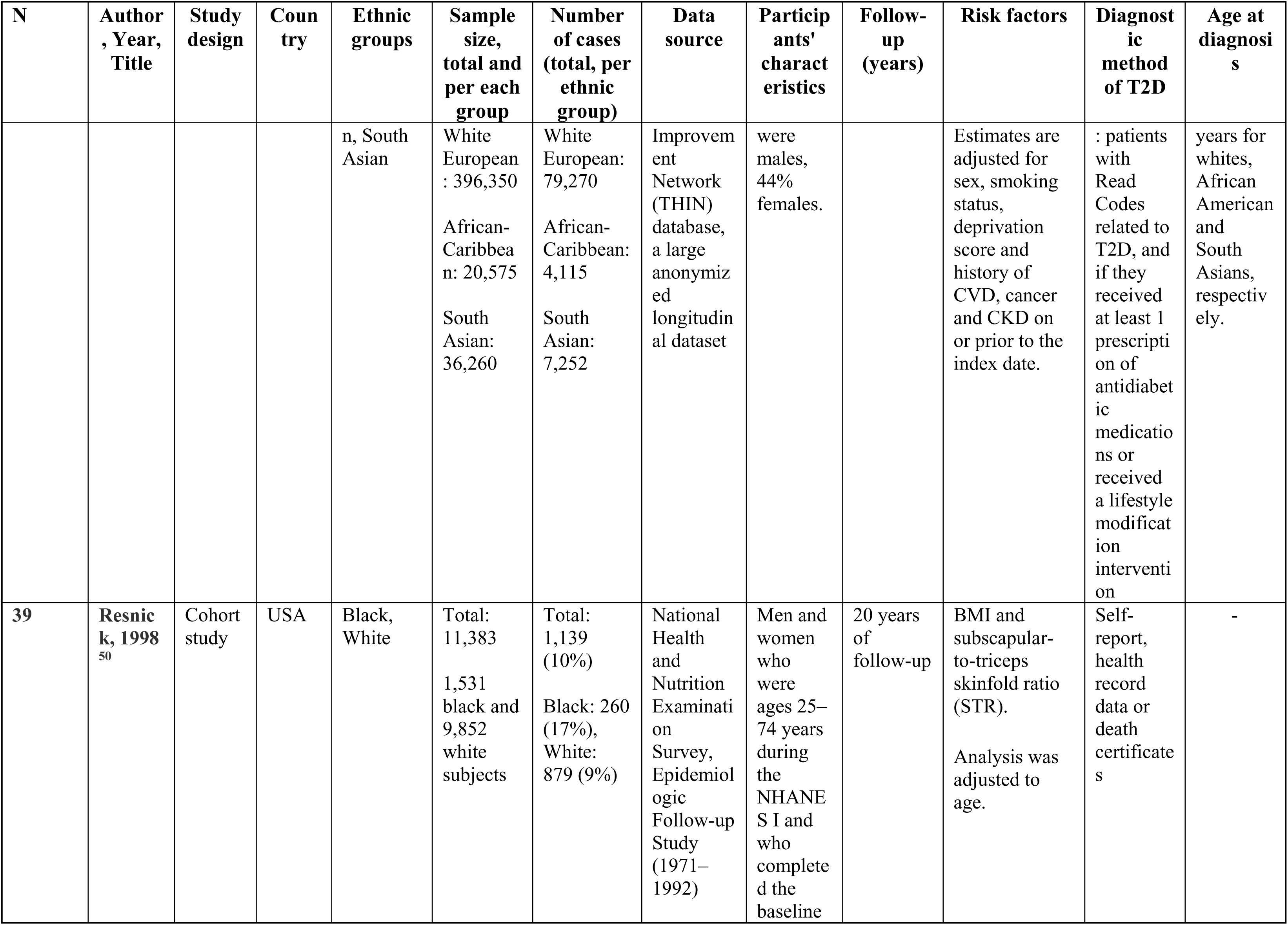
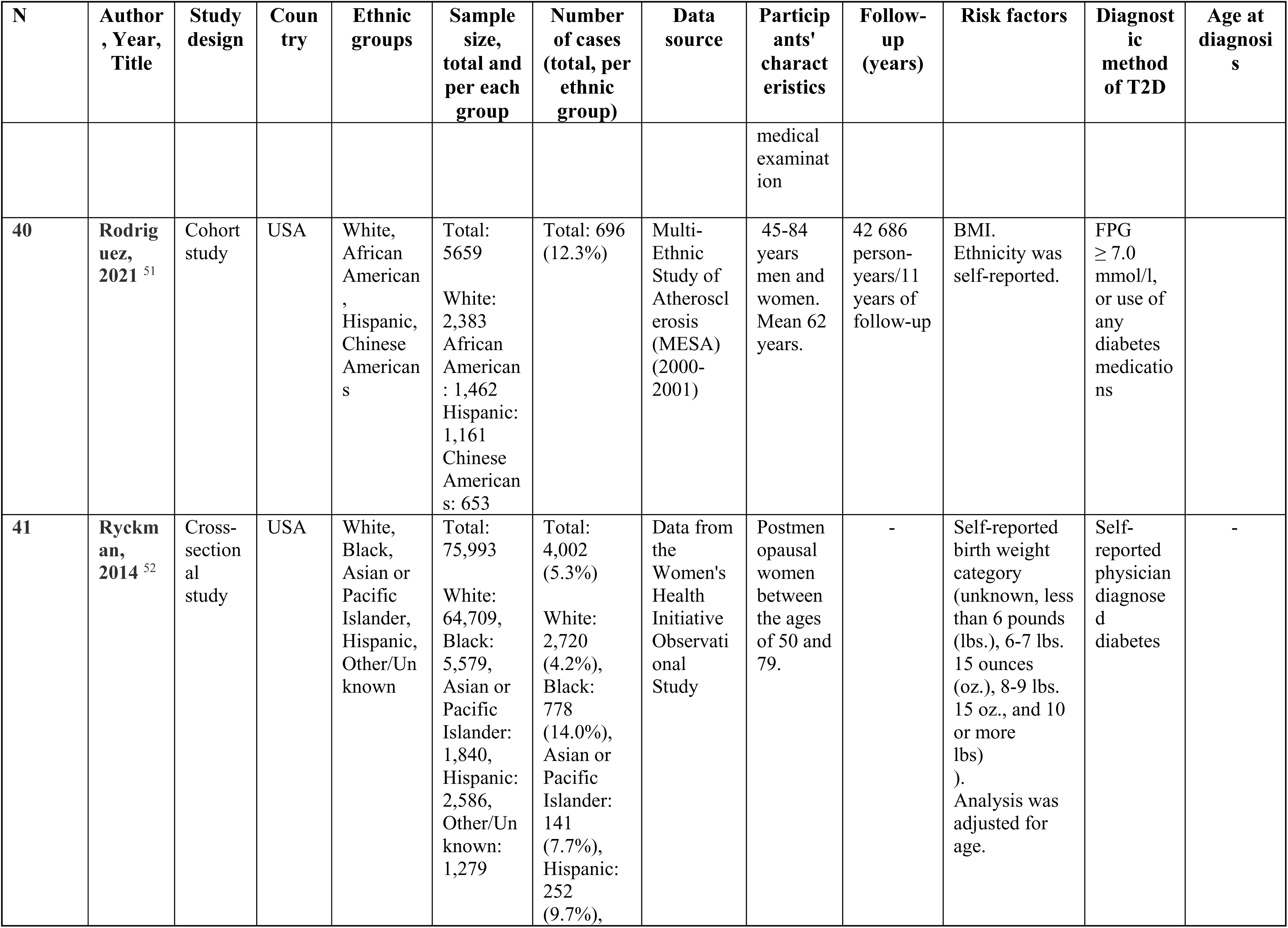
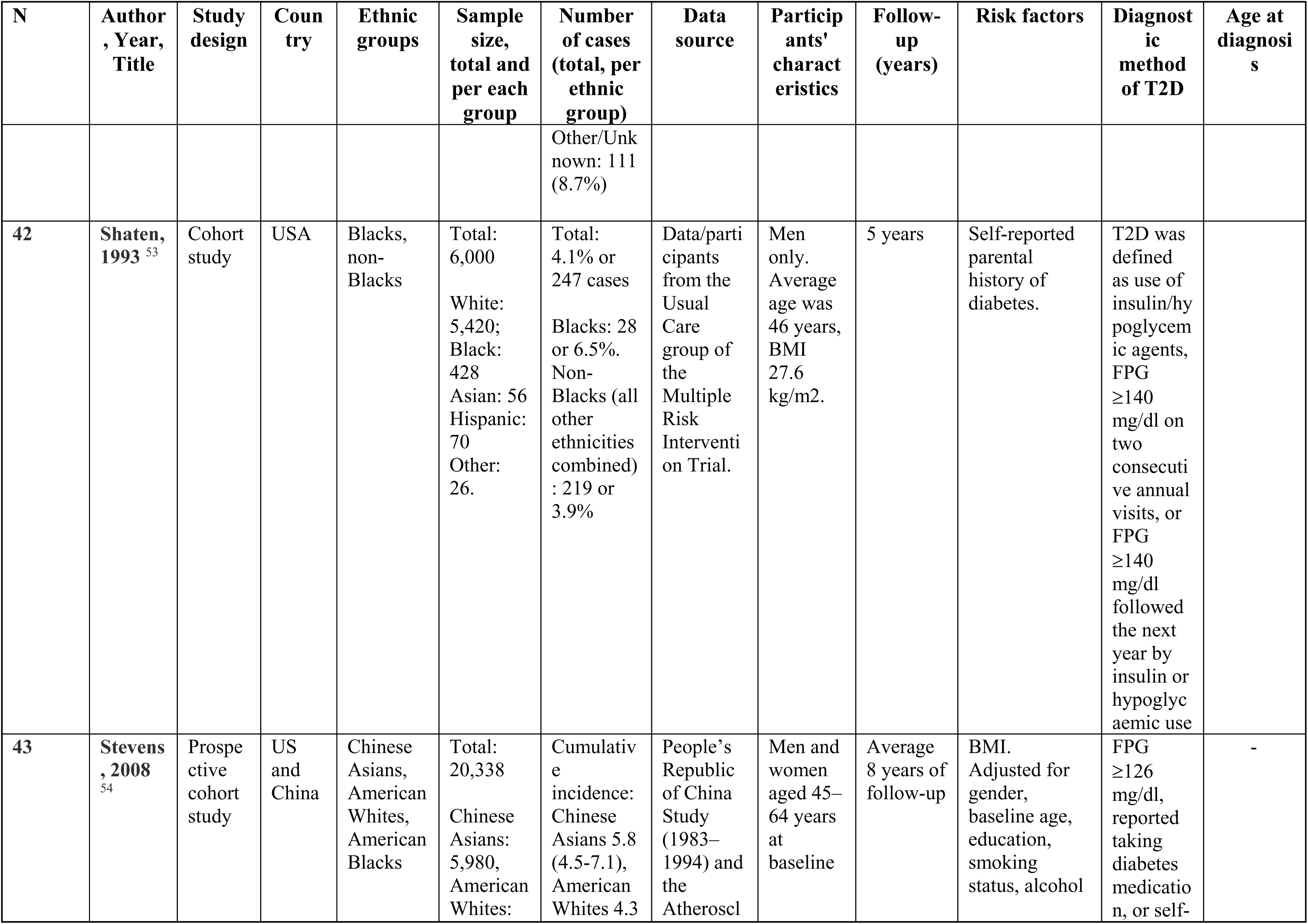
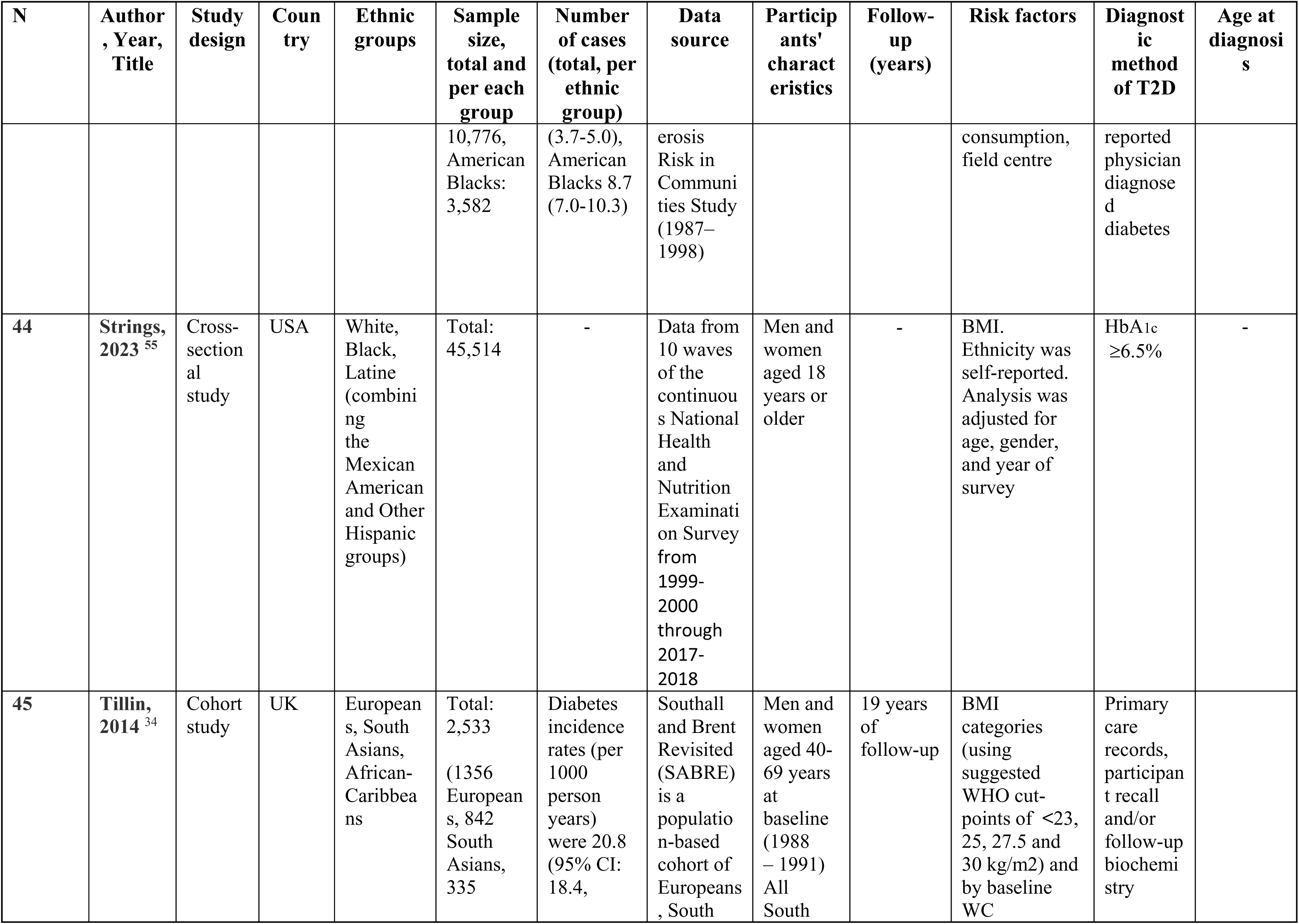
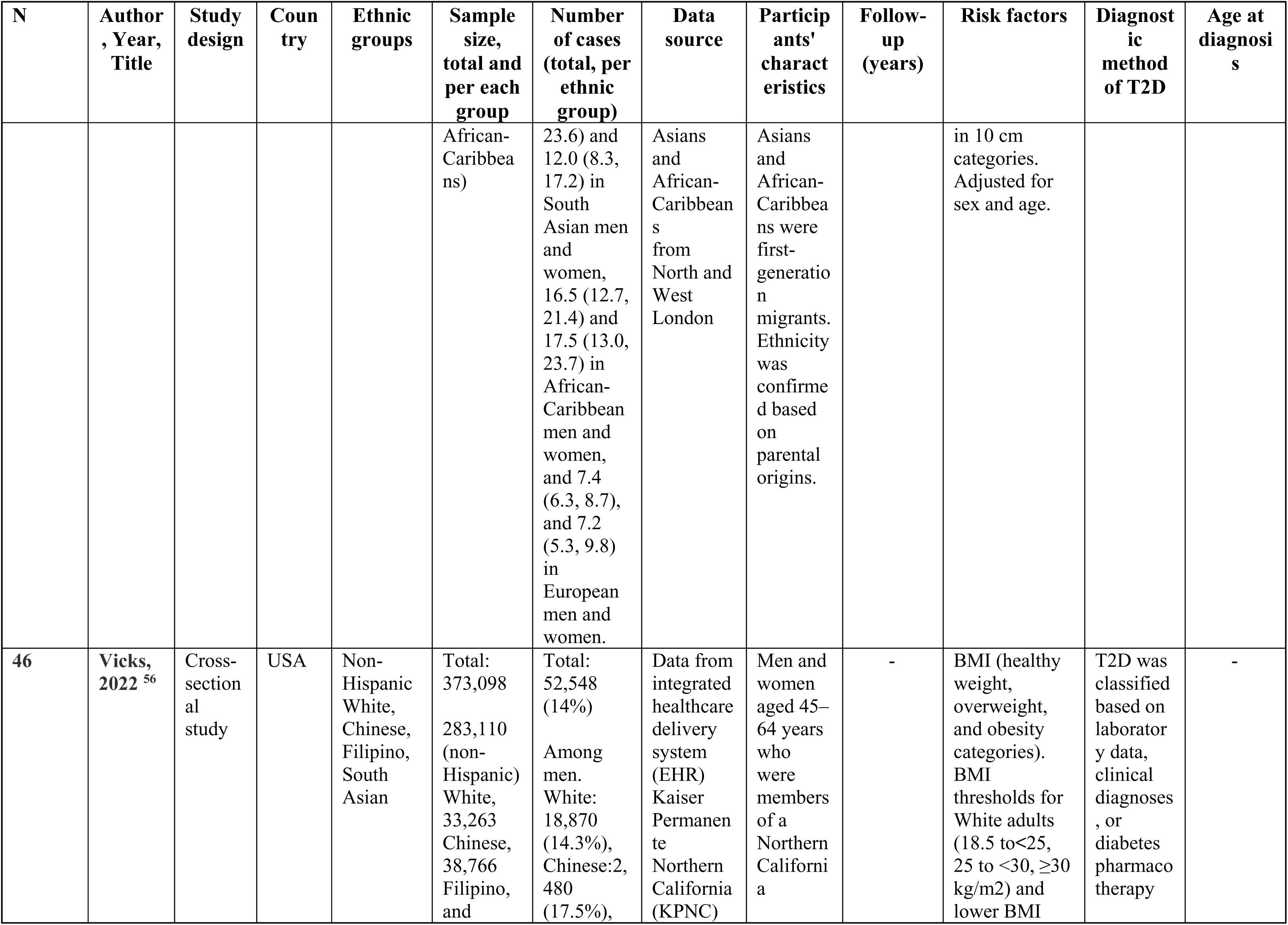
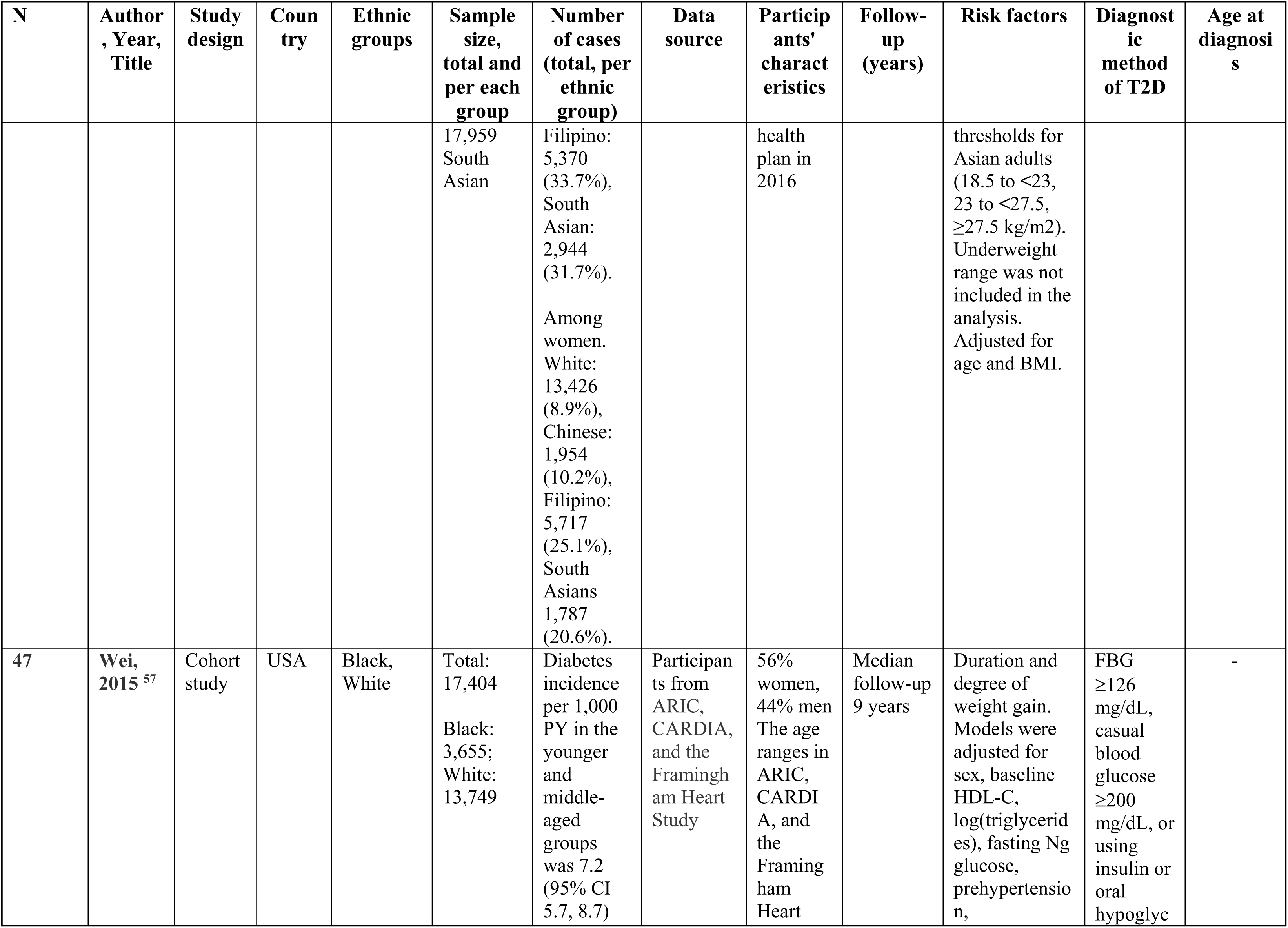
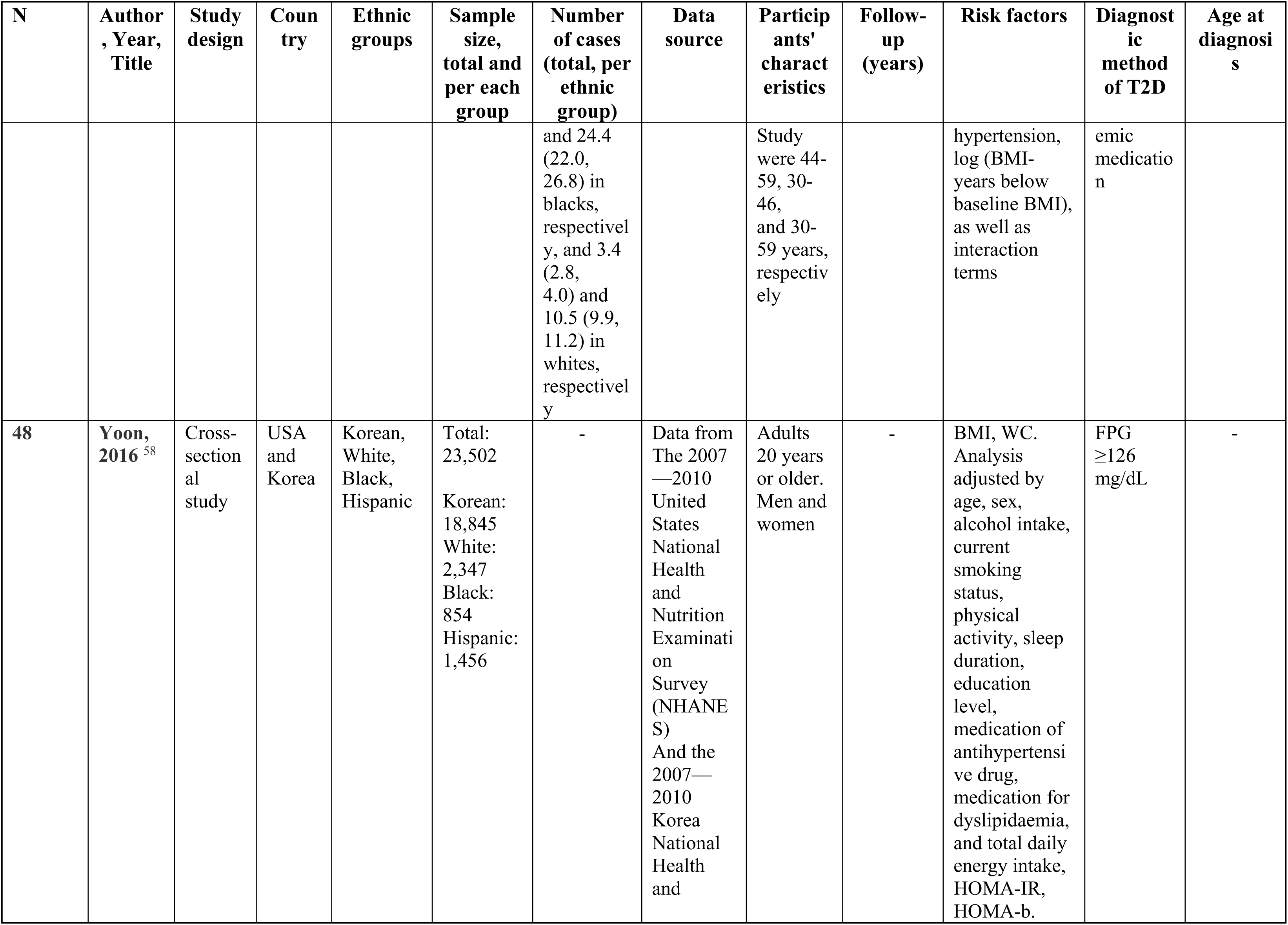
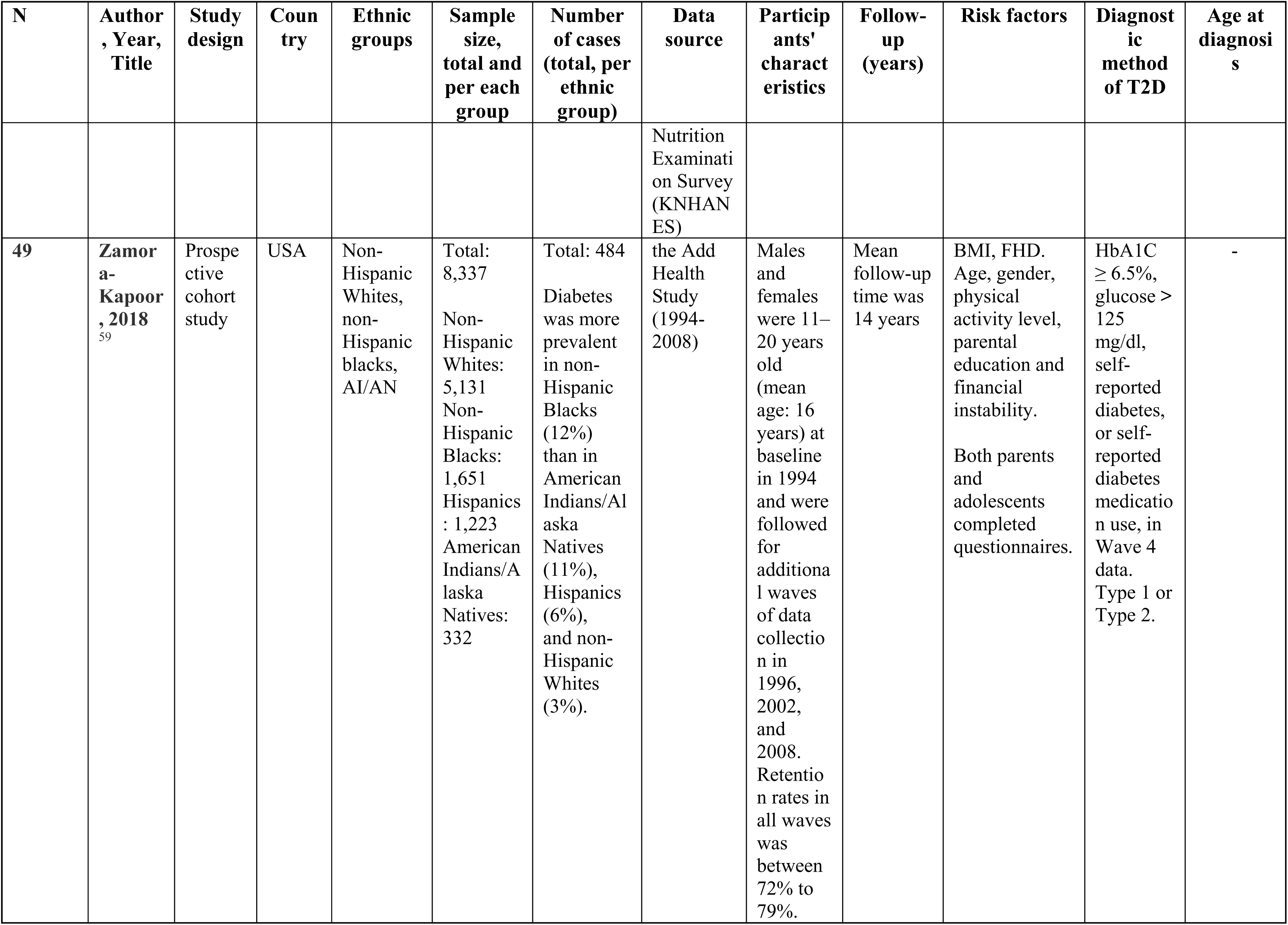
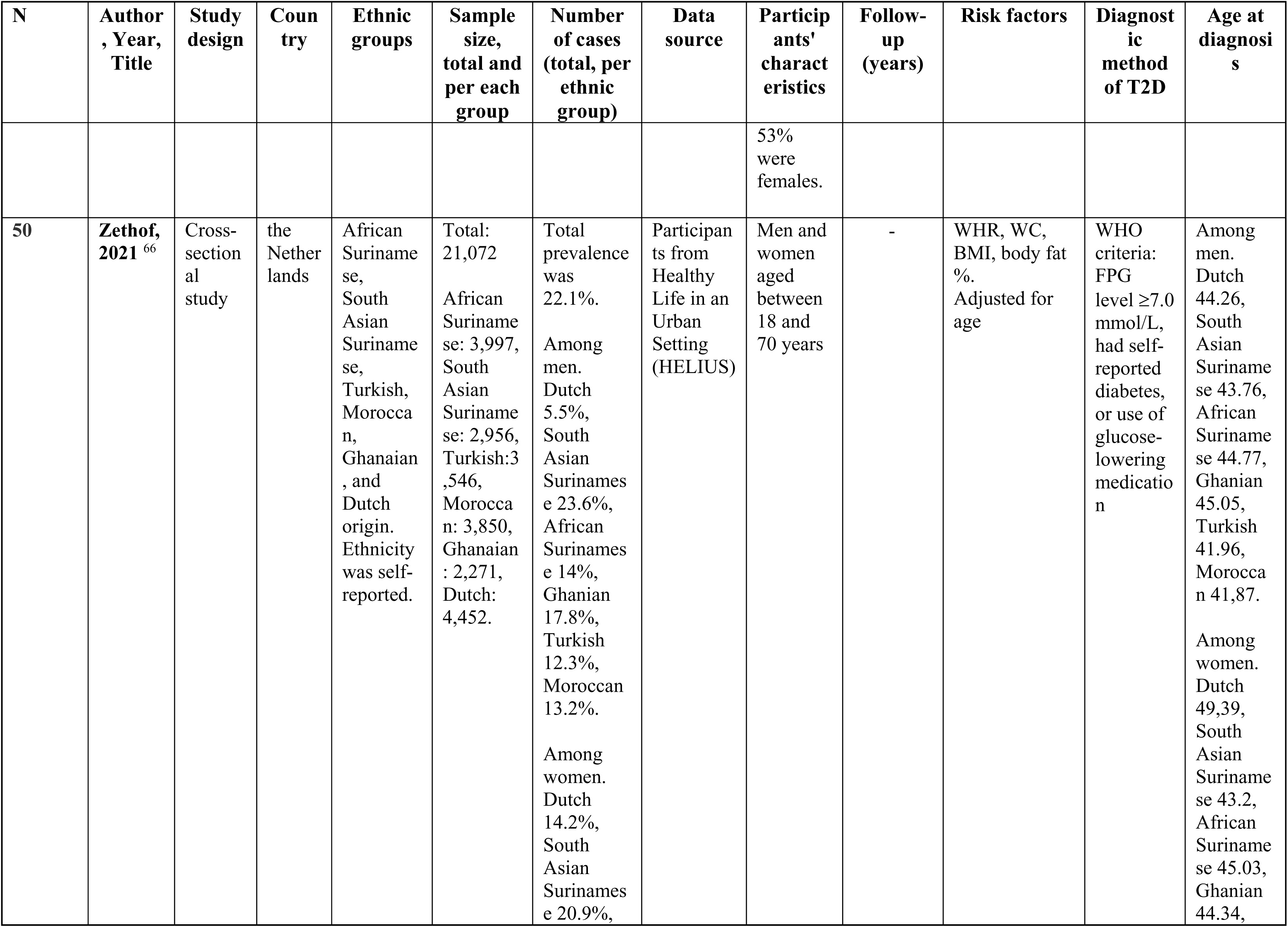
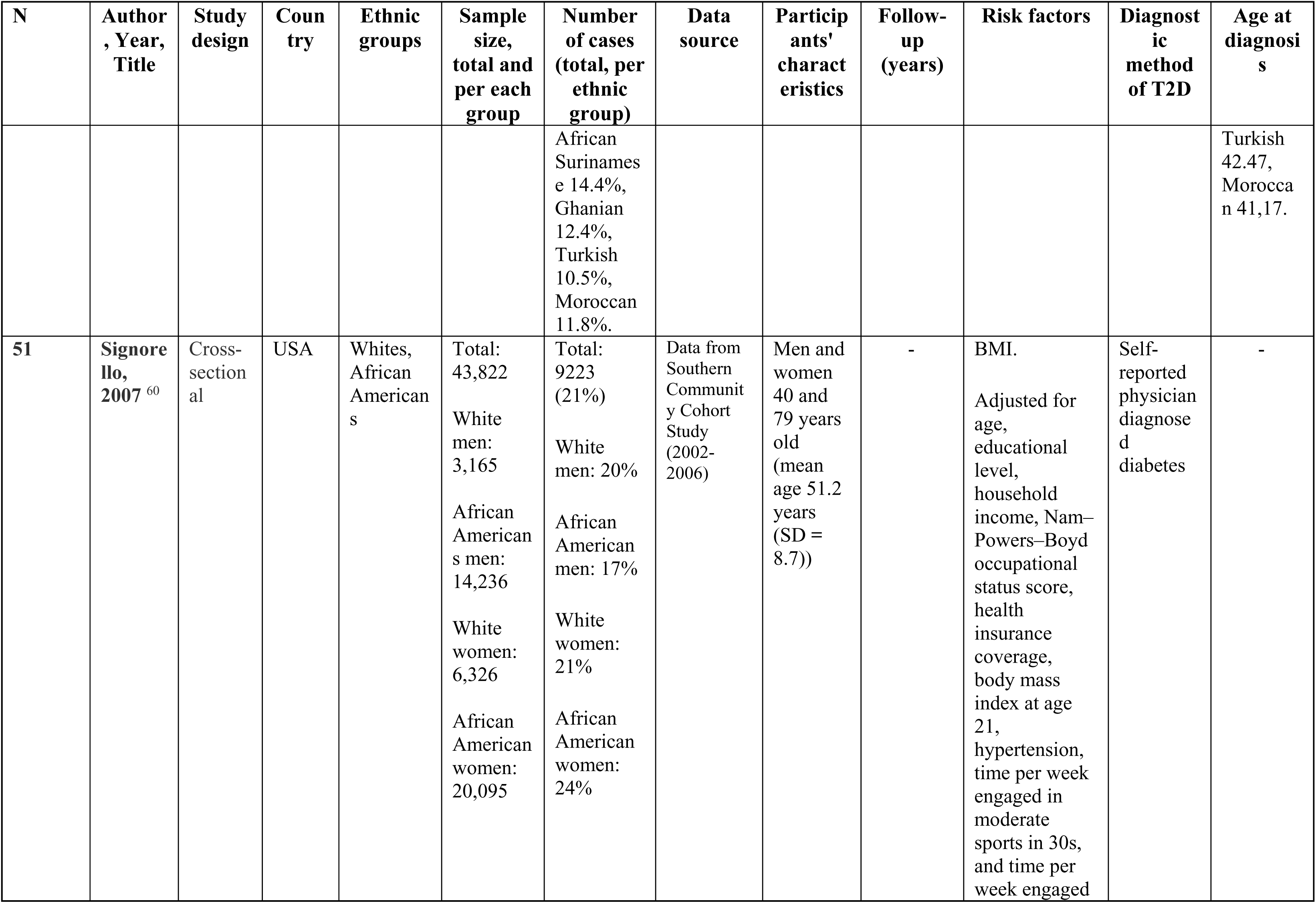
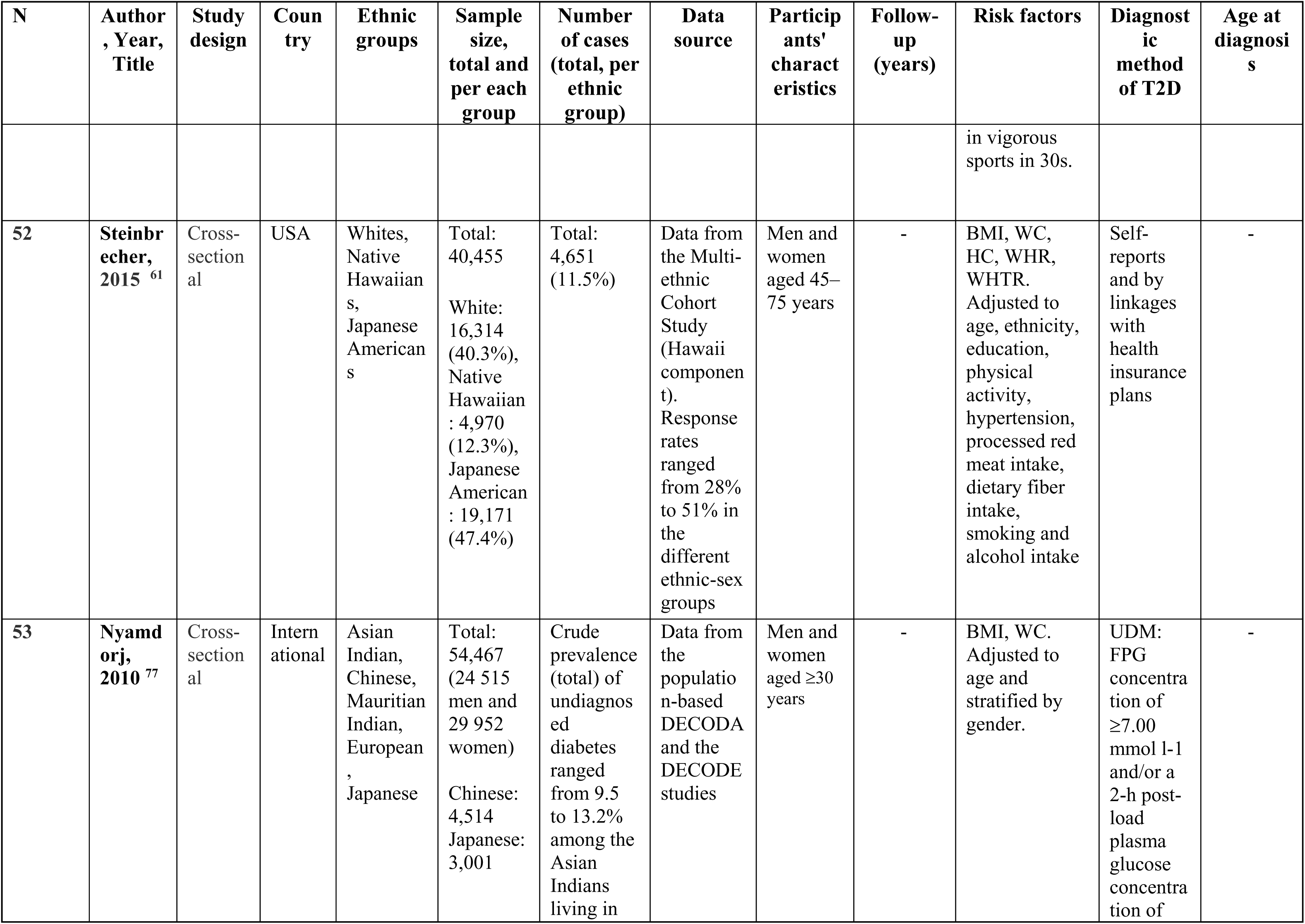
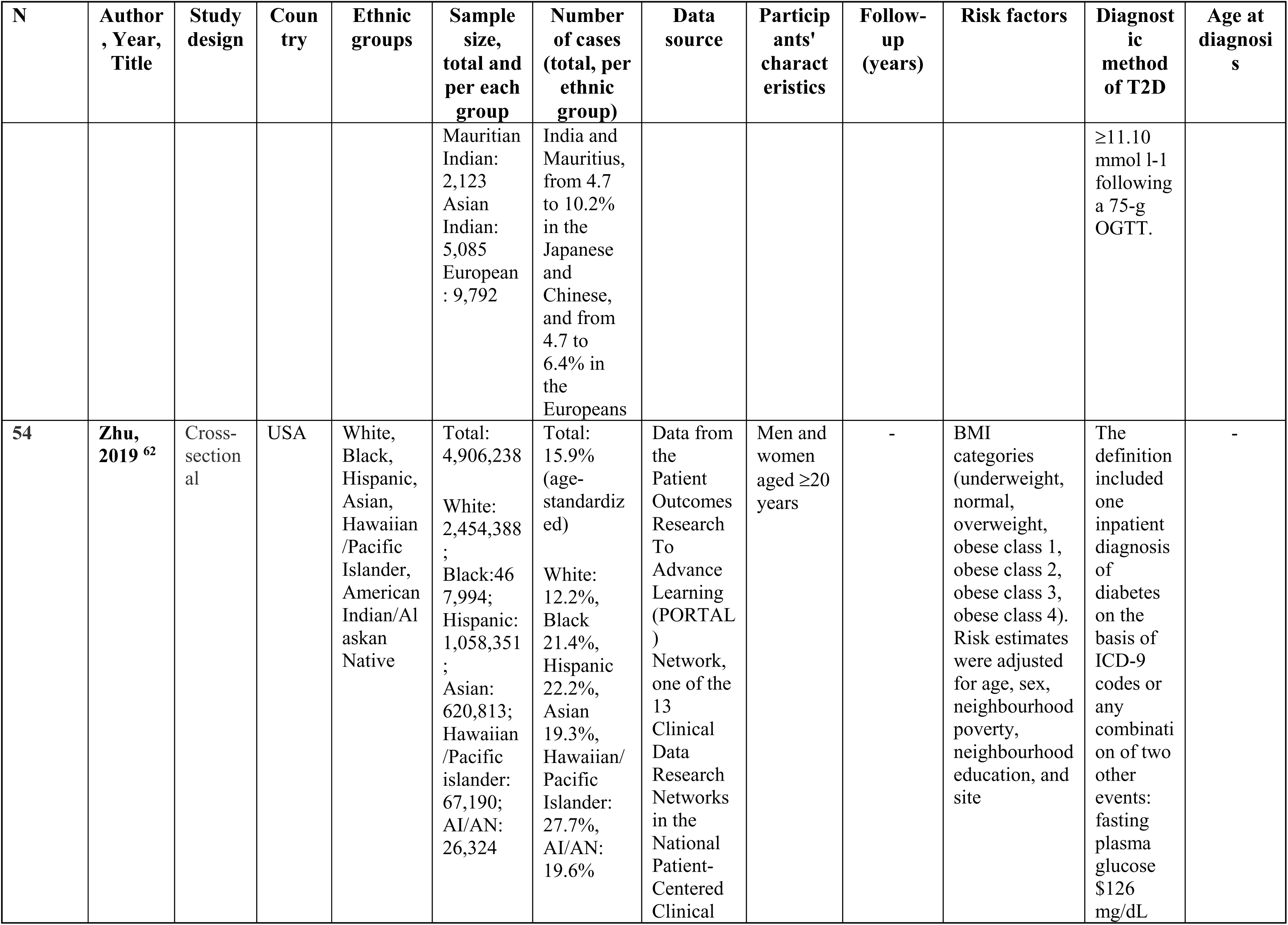
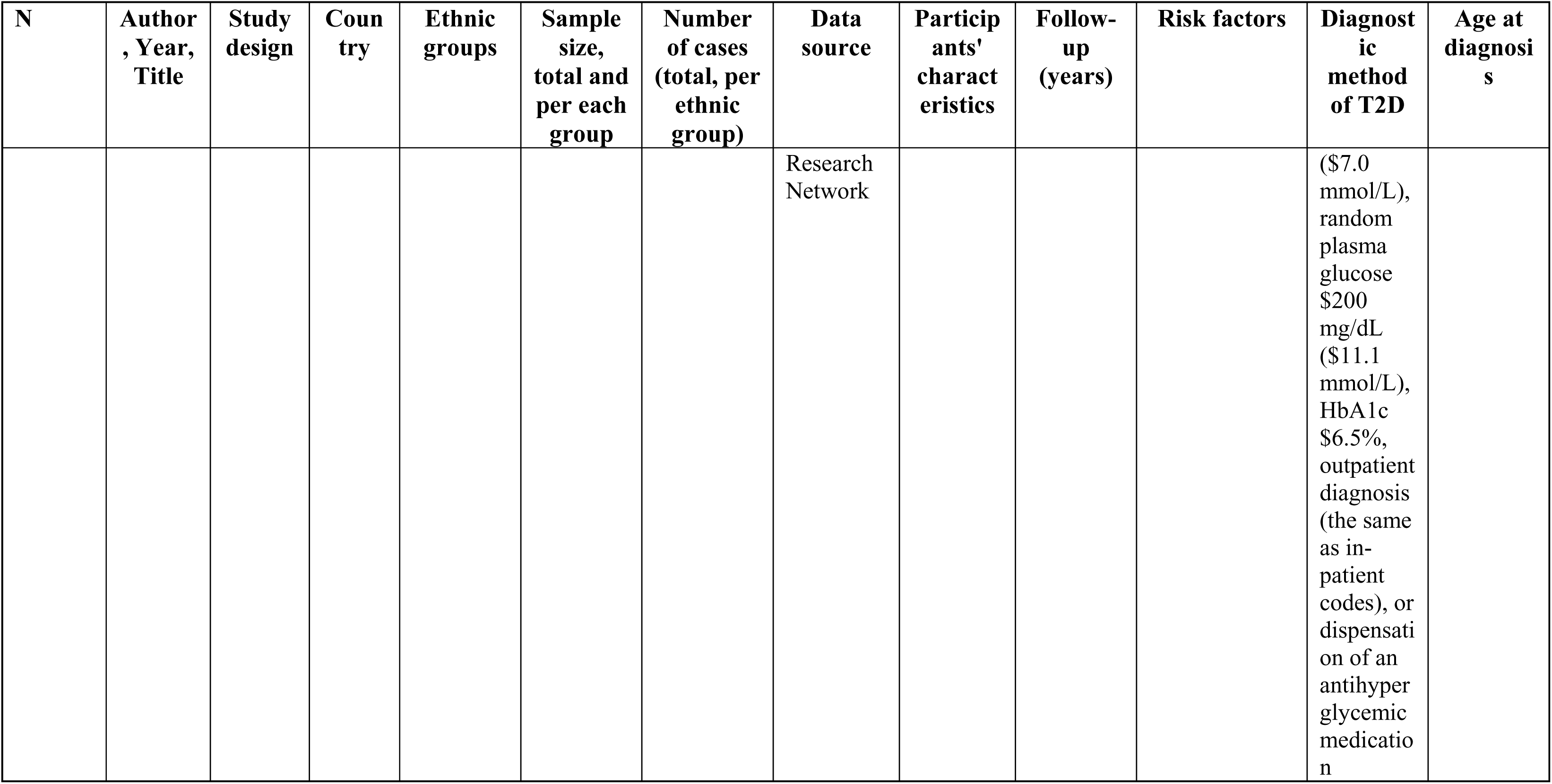
Study characteristics.

The results below focus on anthropometric measures such as BMI, waist circumference (WC), WHR, waist-to-height ratio (WHTR), weight gain, body fat (BF%) percentage (n=50) and non-anthropometric factors related to women’s health (n=5). Results from studies on other factors with less evidence (one study per risk factor) are reported in **Table S3**. The number of studies per anthropometric measure is shown in **Table S4**.

Among case-control studies, three had a moderate risk of bias and one had a low risk of bias. Biases related to high non-response rates differed between cases and controls. Among cohort studies, 12 had a low risk of bias and nine had a moderate risk of bias. Bias was mainly due to differences in the ascertainment of exposure, and the inability to provide evidence that an outcome of interest was not present at the beginning of the study. Among cross-sectional studies, risk of bias was high in seven studies, moderate in eight studies, and low in 14 studies The reasons for potential bias were the absence of standard criteria to measure the risk factor, or the design of the study did not consider and adjust for potential confounding factors. (**Tables S5-7**).

### Descriptive differences across ethnicities in incidence, prevalence, age at diagnosis and anthropometric measures

Prevalence and incidence rates of T2D varied across studies, ethnic groups, and sexes. T2D prevalence was two to four times higher in South Asian, Southeast Asian, Black, and other ethnicities compared to White individuals (8, 14, 28–40). T2D incidence was higher in men among White populations but higher in women among Black populations, with mixed evidence for sex differences amongst South Asians (41) (**Table 1**).

Median age of T2D onset was highest for people of White ethnicity, followed by East Asian, Black, Arab, Southeast Asian and South Asian ethnicities (8, 28, 36). Similarly, the mean age of T2D onset was highest for White individuals (51-58 years), followed by Black (48-54 years), South Asian (44-46 years) and youngest among Turkish and Moroccan individuals (41-42 years) (29, 34, 35, 38, 42) (**Table 1**).

The age-standardized prevalence of T2D in each category of BMI and WC was two to threefold higher in non-white ethnic groups compared to White groups (14, 37, 40, 43–46). Moreover, for any given age of diagnosis, mean BMI was lower for South Asian and Black ethnicities compared with White populations (42).

### Meta-analysis results

#### The relationship between BMI and T2D is strongest in white Europeans and varies across ethnicities

Ethnicity-stratified meta-analysis for the association between BMI and T2D was conducted for people of White, Black and East Asian ethnicities (n=4 studies). People of White ethnicity consistently had the highest odds of T2D within each BMI category compared to people of Black and East Asian ethnicities. When comparing people in obesity category 3 (BMI 40.0-49.9 kg/m^2^) to people with normal weight (BMI 18.5-24.9 kg/m^2^), odds of T2D were increased tenfold for people of White ethnicity (OR 9.95, 95% 5.30-18.66), and fivefold for other ethnic groups (Black OR 5.01,95% 2.57-9.77, East Asian OR 5.11, 95% 4.97-5.26) (**Fig. 2**). (47–50) The included studies had high heterogeneity (I^2^>90%) and most of them a low risk of bias, suggesting the relationship between BMI and T2D could vary across ethnicities. Since only one study reported sex-stratified results, subgroup analyses by sex were not possible.(49) Meta-analysis results of studies reporting hazard ratios (HR, n=2) and relative risks (RR, n=2) per unit increase in BMI are shown in Supplementary materials (**Fig. S1a-b**). The funnel plots showed a possible publication bias (**Fig. S2**). Results from single studies not included in the meta-analysis are described in **Tables S8-9** (n=18 studies).

**Figure 2.**
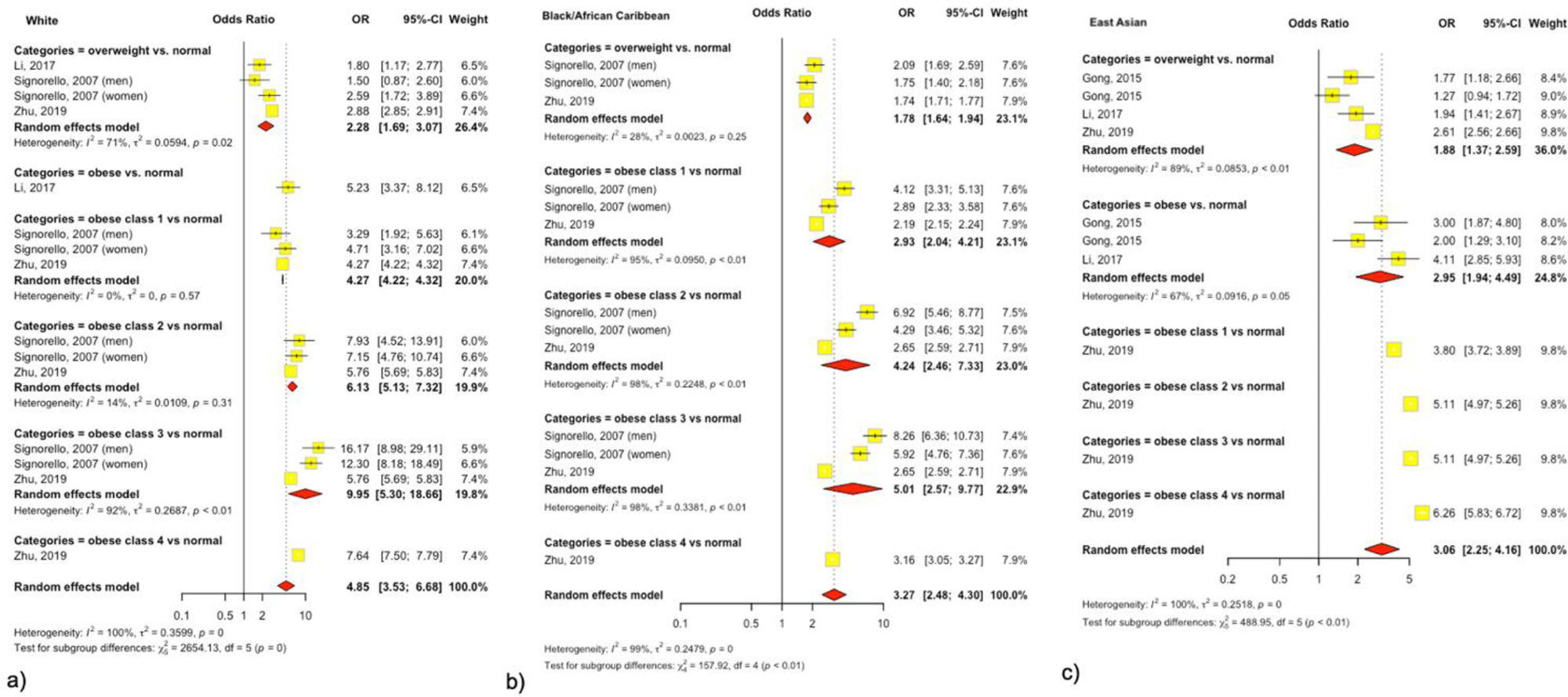
Trans-ethnic meta-analysis of the effect of overweight or obese vs. normal BMI on type 2 diabetes. ((a) White, (b) Black and (c) East Asian ethnic groups)

#### The association of WHR and T2D is stronger in people of Black compared to white ethnicity

Two studies reporting overall and sex-stratified associations between WHR and T2D for people of White and Black ethnicity were meta-analysed.(30, 31) The odds of T2D associated with a one-unit increase in WHR were higher for people of Black ethnicity (OR 2.74, 95% CI 2.22-3.39) compared to people of White ethnicity (OR 2.51, 95% CI 2.30-2.74), with associations being strong among both sexes (**Fig. 3**). Studies for people of White ethnicity were homogeneous (I2=2%), while high heterogeneity was observed for studies including people of Black ethnicity (I2=79%). Most of the included studies had a low risk of bias and potentially publication bias (**Fig. S2**). Results from studies examining the association between WHR and T2D not included in the meta-analysis are described in **Table S10 (n=7 studies)**.

**Figure 3.**
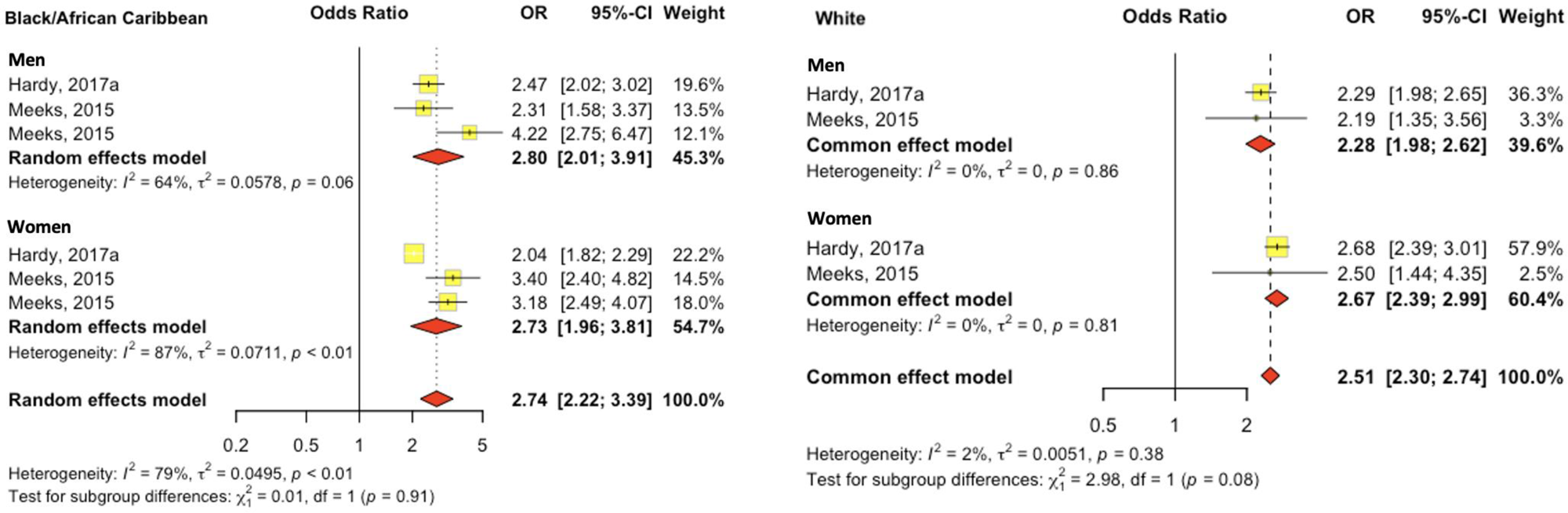
Trans-ethnic meta-analysis of the effect of WHR on type 2 diabetes. (a. Black ethnic group; b. White ethnic group, per unit increase in WHR)

### Narrative synthesis results

#### The effect of WC on T2D differs across ethnicities

Nine studies reported on ethnic differences in the association between WC and T2D, with potential interactions between WC and sex (**Table S11)**. (30, 34, 39, 44, 51–55) Studies reported a stronger association between WC and T2D for East Asian, South Asian, and White individuals than for Hispanic, Native Hawaiian, and Black individuals. (30, 34, 39, 44, 51–55) This trend was more pronounced among women.(44, 56) The Women’s Health Initiative study of postmenopausal women aged 50-79 years observed the strongest association between WC and T2D in East Asians and the weakest in Black women, with American Indian/Alaska Native and Hispanic participants having similar risk to White individuals (53). Despite consistent trends, the overall quality of evidence is low to moderate due to study heterogeneity and bias.

#### WHTR is an independent risk factor for T2D across sexes and ethnic groups

Four studies reported on ethnic differences in the association between WHTR and T2D (**Table S12**). Findings from the Atherosclerosis Risk in Communities (ARIC) (30, 51), the Insulin Resistance Atherosclerosis Study (IRAS) (52) and Multi-Ethnic Cohort (MEC) studies (56) including individuals of White, Black, Hispanic, Japanese and Hawaiian ethnicities demonstrated that WHTR was an independent predictor of T2D across sexes and ethnic groups. In the ARIC (30, 51) and IRAS (52) cohorts, people of White ethnicity had higher odds of T2D associated with WHTR than individuals of Black ethnicity. Despite the low risk of bias in these studies, the evidence for WHTR’s predictive ability across ethnicities is insufficient due to the limited number of studies comparing the same ethnicities and the cross-sectional design of some.

#### The effect of body fat percentage on T2D was stronger among women than men and in White individuals compared to other ethnicities

Four studies reported on ethnic differences in the association between body fat percentage and T2D (**Table S13**). All studies showed a stronger association for White individuals compared to all other ethnic groups, with the association stronger for women compared to men. (31, 38, 52, 53) The IRAS study found this association stronger in White than in Hispanic and Black individuals.(52) The HELIUS study reported a stronger association in White women compared to Black women, with no significant difference among men. (31) Similar trends were seen in a Dutch cross-sectional study(38) and the Women’s Health Initiative cohort(53) , where T2D risk in Black women was weakly associated with BF% but strongly associated with trunk-to-leg ratio (p<0.05 for interaction). The strength of association between T2D and BF % for White (HR 1.38, 95% CI 1.29-1.48) and Hispanic (HR 1.40, 95% CI 1.09-1.80) women was similar (53). Despite consistent trends, the evidence quality was low to moderate due to cross-sectional designs and moderate/high risk of bias.

#### Weight gain is more strongly associated with T2D in white populations

Three studies reported on the association between weight gain and T2D, which was found to be stronger in Japanese and native Hawaiians, followed by individuals of White and Black ethnicities (57–59) Among people of White ethnicity, the association between weight gain and T2D was stronger among women than in men, while the opposite was observed among people of Black ethnicity: for 40+ kg weight gain compared to stable weight (<5kg change) OR 4.0 (CI 3.2-4.9) vs. 3.6 (CI 2.7-4.8) and OR 2.6 (CI 2.3-3.1) vs. 3.3 (CI 2.8-4.0), respectively (57). Similar results were reported in the study which combined ARIC, Coronary Artery Risk Development in Young Adults (CARDIA) and Framingham cohorts, with the association between weight gain and T2D stronger among White individuals than Black subjects: HR per one-unit increment in BMI-years were 1.18 (p=0.02) for younger white individuals, 1.02 (p=0.39) for middle-aged White individuals, 1.35 (p<0.001) in younger Black individuals and 1.11 (p<0.001) in middle-aged Black individuals, respectively (59). However, the limited number of studies and moderate to high risk of bias resulted in low overall evidence quality for this association.

#### The predictive ability of anthropometric measures for T2D and their optimal cut-offs vary across ethnicities

Seven studies assessed the discriminative ability of T2D by anthropometric measures based on the receiver operating characteristics curve (ROC) and the area under the curve (AUC). (30, 36, 51, 52, 55, 60, 61) Measures of central adiposity measures had higher ROC/AUC than overall obesity measures across ethnicities. Five studies suggested ethnicity-specific cut-offs of BMI, WC and WHR to identify high-risk populations for T2D, with lower thresholds for populations of East Asian, South Asian and Black ethnicities compared to people of White ethnicity. (36, 60, 62–64) (**Fig. S3a-c**). The results of other anthropometric measures (skinfold thickness, the ratio of subscapular to triceps, hand grip strength, waist-to-hip-to-height ratio, hip circumference, a body shape index, body adiposity index, body height, trunk fat and trunk-to-leg ratio) with less evidence are shown in the **Table S14**. Due to the high proportion of cross-sectional studies and moderate/high risk of bias, these results have low/moderate evidence quality and should be interpreted cautiously.

#### Women-health associated factors

We found per one study for pregnancy index (65), breastfeeding (66), birth weight (67), and age at menarche (29). Ethnic differences were noted in the relationships between pregnancy index, birth weight, age at menarche, and T2D, but not for breastfeeding, which was protective for all ethnic groups (**Table S15**). After adjustment for different confounders, a higher index of parity was associated with an elevated risk of T2D among White, South Asian and Chinese women, but Chinese women had a higher risk of T2D for 3-4 (13%) and ≥5 (359%) deliveries compared to women with 1 delivery than South Asian (3% and 23%) and White (13% and 52%) women during almost 12 years of follow-up.(65) Higher birth weight increased T2D odds for East Asian women but decreased it for White, Black, and Hispanic women (67). Associations between breastfeeding, age at menarche, and T2D were non-significant in adjusted models (29, 66). The quality of evidence for studies examining women’s health related factors was low.

The results for FHD are shown in **Fig. S4 and Table S16.**

## Discussion

This systematic review of 54 studies and meta-analysis of 12 studies summarized evidence on ethnic differences in the association between anthropometric measures, women-health-related factors, and T2D. Consistent with previous research, ethnic- and sex-differences in T2D risk were evident, and not always in a consistent direction. The median age of T2D diagnosis was 10 years earlier for South Asian individuals, seven years earlier for Black individuals, and five years earlier for East Asian compared to White individuals, suggesting that people of diverse ethnic backgrounds might benefit from earlier screening (<40 years) (14).

Our synthesis suggests that while BMI and weight gain are strongly associated with T2D in White groups, these measures are poorer predictors of T2D for people of South Asian, East Asian and Black ethnicities. Furthermore, we find that ratio measures of central adiposity such as WHR, and WHTR, may better predict T2D in all ethnic groups and both sexes, especially for people of Black ethnicity, for whom other anthropometric measures were less effective. BF % and WC were better predictors of T2D in women than in men, particularly those of White, Hispanic and South Asian ethnicities, but less so for those of Black ethnicity. However, WC’s effectiveness is influenced by height (36), making WHR and WHTR more reliable. As individuals’ height is relatively stable during adulthood, WHTR could better capture changes in WC, but our review identified fewer studies comparing its effect across ethnicities.

It is known that BMI does not reflect the proportions of lean and fat mass, where the contribution of the latter is more important in T2D development.(36) Moreover, it does not differentiate subcutaneous (SAT), visceral (VAT) adipose tissues and ectopic fat distributions, which are stronger associated with insulin resistance than general adiposity (52, 68). Previous findings found that Asian individuals have more VAT and ectopic fat than people of White ethnicity, while people of Black ethnicity have less VAT compared to people of Asian and White ethnicities after adjustment to various total adiposity measures (52, 54, 69, 70). Higher VAT is associated with enhanced proinflammatory markers and greater liver fat deposition leading to insulin resistance (71). It was previously proposed that South Asians may have less capacity to store VAT at the same BMI levels, enhancing fat deposition in secondary depots such as the liver and pancreas (70, 72). That means that changes in VAT may have more deteriorating effects on this group compared to white Europeans. However, despite having less VAT after controlling for BMI, people of Black ethnicity remained at a greater risk for T2D compared to those of White background (70). This means different ethnicities may have distinct pathophysiology of T2D and contributing genetic susceptibility (33, 54). It is still unclear if these ethnic differences occur due to variations in genetics, biological characteristics or lifestyle factors (8).

Therefore, relying solely on BMI might fail to identify high-risk ethnic groups or make weight-loss interventions ineffective for those with lower BMI but higher visceral fat. On the contrary, ratio measures of central adiposity may better represent changes in abdominal fat distribution, which are more strongly linked to insulin resistance (60). Currently, the US Preventive Services Task Force (USPSTF) and American Diabetes Association (ADA) recommendations for earlier initiation of screening for T2D (>35 years and any age) are mainly based on selecting individuals with overweight and obese BMI categories (15, 16). Therefore, these guidelines may miss individuals of diverse backgrounds for whom BMI does not reflect T2D risk. Decreasing the T2D screening age for populations of diverse backgrounds and incorporating central adiposity measures into clinical practice could help to decrease health inequalities and improve timely preventive interventions. Since the quality of evidence coming predominantly from cross-sectional studies is low, more longitudinal research with a bigger population of younger ages studying the effect of changes in central obesity measures across ethnicities is needed.

This review demonstrated an emerging significance of sex-specific risk factors among women, even though the quality of evidence was low. East and South Asian women aged 18-50 had a higher T2D risk with increasing parity than White women in a Canadian study, with associations between parity and T2D strongest for Chinese women followed by South Asian and White women. Possible mechanisms could be mediated effect via postpartum weight retention and increased adiposity (65, 73). However, a sensitivity analysis in this study showed that the attenuated but still significant effect remains even after adjustment for BMI, meaning parity may be an independent risk factor for diabetes. Ethnic differences in the effect of parity could be associated with differences in genetics. Previous studies demonstrated that Chinese women may have lower adiponectin levels in pregnancy, which is associated with postpartum insulin resistance and B-cell dysfunction (74, 75) and an increased risk of metabolic syndrome (76), while women of South Asian ethnicity may have other much stronger risk factors leading to an increased risk of T2D (65). Another study revealed that after taking into account the histories of parity and breastfeeding, women with ≥5 parities had elevated T2D risk, regardless of breastfeeding duration, but adjusting for weight gain women with ≥3 parities did not have higher diabetes risk when they breastfed more than 12 months in total or each child more than 3 months (53). Breastfeeding’s benefits might include higher energy expenditure and enhanced insulin sensitivity. Overall, breastfeeding was associated with reduced T2D risk across different ethnicities. Therefore, women with high parity potentially could be a target population for the prevention of T2D. More longitudinal studies which control for socio-economic variables and adiposity measures are needed to examine ethnic differences in the relationship between age at menarche, birth weight and T2D.

## Strengths and limitations

As only studies published in English were included, there is a potential for some findings from other non-English publications to be missed, which may result in a lack of findings from certain countries. However, it is difficult to predict how it might influence the review results. Secondly, we did not include studies focusing on environmental, clinical, genetic and lifestyle factors such as diet and physical activity which were out of the scope of this review, but which still contribute to the development of diabetes. On the other hand, this systematic review has several strengths. Firstly, we did not restrict the search to certain clinical risk factors which allowed us to capture a broader number of studies. Secondly, the search strategy allowed us to identify evidence of ethnic differences in the effect of anthropometrics and the emerging importance of women-health-related factors and T2D for all ethnicities. In addition, this narrative synthesis allowed us to highlight the ethnic heterogeneity in the predictive ability of T2D using anthropometric measures, which are not currently considered in global multi-ethnic populations.

## Conclusion

This systematic review showed that T2D affects disproportionately populations from different ethnic backgrounds at significantly younger ages. Our results demonstrated that ratio measures of central adiposity such as WHR, or potentially WHTR, could be better at identifying high-risk populations for T2D across ethnicities and sexes than widely-used BMI, with the importance of stratification of Asian ethnicities when assessing T2D risk, however, most of the quality of evidence is moderate/low. Additionally, this systematic review highlighted the importance of ethnic- and sex-specific risk factors such as number of parities and breastfeeding among women. These factors should be considered when assessing the risk of diabetes among women, especially those from non-White ethnic backgrounds, which could be a target population for prevention. More longitudinal studies with representative populations are needed to explain these mechanisms.

## Data Availability

All data produced in the present work are contained in the manuscript

## Contributors

BO, RM, SF and MKS developed the idea and study design. BO and TH did the data search, extraction and quality assessment, with the input from RM, SF and MKS. BO did the meta-analysis. BO wrote the first version of the manuscript with the substantial contributions from all authors. SH, MS, DS, MKS and MS contributed to the interpretation of findings and critical revision of the manuscript. All authors approved the final version of the manuscript for submission.

## Declaration of interests

We declare no competing interests.

## Acknowledgements

The authors thank Paula Funnel, the Faculty Liaison Librarian for Medicine & Dentistry at Queen Mary University of London, for assisting with the definition and validation of keywords and MESH terms.

## Notes

### Competing Interest Statement

The authors have declared no competing interest.

