## Supplementary Tables and Figures for "Evidence of ethnic variations in the relationships between routinely recorded clinical factors and T2D: a systematic review and meta-analysis"

### Supplementary materials, methods and results

**Table S1.** MESH terms and keywords used in the Medline Complete database.

| | |
|---|---|
| **Medline** | TI ((MH "Diabetes Mellitus, Type 2+") OR (MH "Diabetes Mellitus+") OR "type 2 diabetes" OR "type II diabetes" OR "diabetes mellitus") AND TI ("ethnicity" OR "Ethnic minority" OR "Multi-ethnic" OR "Ethnic" OR (MH "Ethnic and Racial Minorities") OR (MH "Ethnicity") OR "race-ethnic" OR "race" OR "Chinese" OR "Arab" OR "Asian" OR "African" OR "Caribbean" OR "Black").<br>Filters applied: human, English, 1990-2023, Journal article, Medline database |

**Table S2**. Categorisation of ethnicities from included studies into higher-level ethnicities

| | |
|---|---|
| **South Asian** | • Indian/ American Indian/ Asian Indian/ Hindustani Surinamese<br>• Pakistani<br>• Bangladeshi<br>• Sri Lankan<br>• South Asian/ South Asian Surinamese |
| **Black** | • Black/ Non-Hispanic Black/ Black American/ African-Caribbean/ African Surinamese<br>• Jamaican<br>• Nigerian<br>• Mauritius Creoles<br>• Ghanian |
| **East Asian** | • East Asian<br>• Korean<br>• Japanese/ Japanese American<br>• Chinese/ Han Chinese/ Chinese American |

| | |
|---|---|
| | • Uyghur |
| **Southeast Asian** | • Filipino<br>• Vietnamese<br>• Malay |
| **White** | • White/ US White/ UK White/ Non-Hispanic White<br>• European<br>• Norway<br>• Dutch |
| **Hispanic** | • Hispanics/ Hispanic American<br>• Mexican/ Mexican American<br>• Latino/ Latin American |
| **Middle East/ Arab** | • Arab<br>• United Arab Emirates<br>• Iraq<br>• Moroccan<br>• Turkish |
| **Others** | • Hawaiian<br>• Aboriginal<br>• Other Indigenous<br>• American Indians/Alaska Natives<br>• Pacific Islanders<br>• Mauritian<br>• Asian/Asian American |

**Table S3**. A table with results for five single risk factor studies (Supplementary excel file)

**Table S4**. Number of studies per anthropometric measures and unit measures.

| Anthropometric measure | Number of studies considered | Ethnicities represented |
|---|---|---|
| **BMI (categorical)** | 9 | South Asian, East Asian, White, Others, American Indian, Black/African Caribbean, Hispanic |

| | | |
|---|---|---|
| **BMI (per SD or linear)** | 15 | Black/African Caribbean, White, Hispanic, East Asian, South Asian, American Indian, Others |
| **Weight gain (categorical)** | 2 | White, Black/African Caribbean, Hawaiian, Japanese American |
| **Weight gain (per 1 log-increase in BMI-year)** | 1 | White, Black/African Caribbean |
| **Body fat percentage (per SD or linear)** | 5 | Black/African Caribbean, White, Hispanic, South Asian, East Asian, Turkish, Moroccan |
| **Trunk fat, trunk-to-leg-ratio (per quartile and linear/unit)** | 1 | Black/African Caribbean, Hispanic |
| **WC (per SD or linear)** | 9 | Asian Indian, Mauritian Indian, East Asian, White, Iraqi, Black/African Caribbean, Turkish, Southeast Asian, Hispanic, Others |
| **WC (categorical)** | 1 | Jamaican, U.S. |
| **WHR (per SD or linear)** | 11 | South Asian, East Asian, Southeast Asian, White, Black/African Caribbean, Hispanic, Native Hawaiian, Japanese American, Turkish, Moroccan, Others |
| **WHTR (per SD and linear)** | 4 | Black/African Caribbean, White, Hispanic, Native Hawaiian, Japanese |
| **WHHR (per unit/linear)** | 1 | White, Black/African Caribbean |
| **Body height (per unit/linear [cm])** | 1 | White, South Asian |
| **Skinfold thickness/Triceps skinfold/Subscapular skinfold/Sum of skinfold thickness (per SD or linear)** | 2 | White, Hispanic, Black/African Caribbean/African American |
| **The ratio of subscapular to triceps (per unit/linear)** | 2 | White, Hispanic, Black/African Caribbean/African American |
| **Hand grip strength (per SD)** | 1 | White, Black/African Caribbean, South Asian |
| **Hip circumference (per SD)** | 2 | White, Native Hawaiian, Japanese American, Black/African Caribbean, Hispanic |

| | | |
|---|---|---|
| **ABSI (per SD)** | 2 | White, Black/African Caribbean |
| **BAI (per SD)** | 2 | White, Black/African Caribbean |
| **Birth weight (categorical)** | 1 | White, Black/African Caribbean, Asian/Pacific Islander, Hispanic |
| **Combination of anthropometric measures** | 2 | White, Native Hawaiian, Japanese American, Black, Aboriginal, South Asian, Hispanic |

**Table S5**. Risk of bias assessment. NOS scale for case-control studies

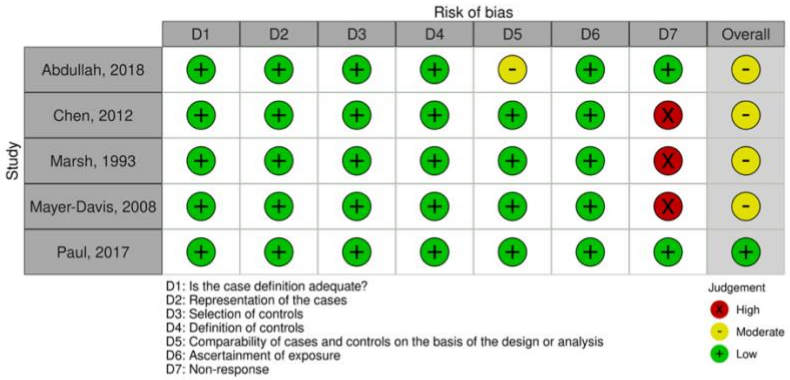


.

**Table S6.** Risk of bias assessment. NOS scale for cohort studies.

| Study | D1 | D2 | D3 | D4 | D5 | D6 | D7 | Overall |
|---|---|---|---|---|---|---|---|---|
| Caleyachetty, 2021 | + | + | + | + | + | + | + | + |
| Dreydus, 2012 | + | + | + | + | + | + | + | + |
| Hardy, 2017 | + | + | + | + | + | + | + | + |
| Jeong, 2021 | + | + | + | + | + | + | + | + |
| Mackay, 2009 | + | + | + | + | + | + | + | + |
| Almahmeed, 2017 | + | + | + | + | + | + | + | + |
| Chan, 2018 | + | + | + | + | + | + | + | + |
| Chatterjee, 2016 | + | + | + | + | + | - | + | - |
| Chiu, 2011 | + | + | X | + | + | + | + | - |
| Gurk, 2017 | + | + | + | X | + | + | + | - |
| Kulick, 2016 | + | + | + | X | + | + | + | - |
| Luo, 2019 | + | + | + | + | + | + | + | + |
| Lutsey, 2010 | + | + | + | + | + | + | + | + |
| Ma, 2012 | + | + | + | + | + | + | + | + |
| Maskarinec, 2009 | + | + | + | + | - | + | + | - |
| Morimoto, 2011 | + | + | + | + | - | + | + | - |
| Nan, 2008 | + | + | + | + | + | + | + | + |
| Narayan, 2021 | + | + | + | + | + | + | + | + |
| Piccolo, 2014 | + | + | + | + | + | - | + | - |
| Piccolo, 2013 | X | + | + | + | + | + | + | - |
| Reis, 2016 | + | + | + | + | + | + | + | + |
| Resnick, 1998 | + | + | + | - | + | + | + | - |
| Rodriguez, 2021 | + | + | + | - | + | + | + | - |
| Shaten, 1993 | + | + | + | + | + | + | + | + |
| Stevens, 2008 | + | + | - | + | + | + | + | - |
| Tillin, 2013 | + | + | + | + | + | + | + | + |
| Tillin, 2014 | + | + | + | + | + | - | + | - |
| Wei, 2011 | + | + | + | + | + | + | + | + |
| Wei, 2015 | + | + | + | + | + | + | + | + |
| Zamora-Kapoor, 2018 | + | + | + | + | + | + | + | + |
| Beihl, 2009 | + | + | + | + | + | + | + | + |

D1: Representativeness of the exposed cohort
D2: Selection of the non-exposed cohort
D3: Ascertainment of the exposure
D4: Demonstration that outcome of interest was not present at the start of study
D5: Comparability of the cohorts on the basis of the design or analysis
D6: Ascertainment of exposure
D7: Non-response rate

Judgement: X High; - Moderate; + Low

**Table S7.** JBI critical appraisal checklist for cross-sectional studies

| Study | D1 | D2 | D3 | D4 | D5 | D6 | D7 | D8 | Overall |
|---|---|---|---|---|---|---|---|---|---|
| Alperet, 2016 | + | + | X | + | X | X | + | - | X |
| De Koning, 2010 | + | + | + | + | + | + | + | + | + |
| Hardy, 2017 | + | + | + | + | + | + | + | + | + |
| Meeks, 2015 | + | + | + | + | X | - | + | + | - |
| Aggarwal, 2022 | + | + | + | + | X | - | + | + | - |
| Bennet, 2014 | + | + | + | + | + | + | + | + | + |
| Cheong, 2014 | + | + | + | + | X | - | + | + | - |
| Cheong, 2013 | + | + | + | + | X | - | + | + | - |
| Cohen, 2009 | ? | + | X | X | + | + | X | + | X |
| Diaz, 2007 | + | + | + | + | X | - | + | + | - |
| Gong, 2015 | + | + | + | + | + | + | + | + | + |
| Hamoudi, 2019 | + | + | + | + | X | - | + | X | X |
| Huxley, 2008 | X | ? | + | + | X | - | + | + | X |
| Jenum, 2012 | + | + | + | + | + | + | + | + | + |
| Jenum, 2005 | + | + | + | + | + | + | + | + | + |
| Li, 2017 | + | + | X | X | + | + | - | + | X |
| Lorenzo, 2007 | + | X | + | + | X | - | + | + | X |
| Ntuk, 2017 | + | + | + | + | + | + | + | + | + |
| Ntuk, 2014 | + | + | + | + | + | + | + | + | + |
| Okosun, 1998 | + | + | + | + | X | X | + | + | - |
| Ryckman, 2014 | + | + | X | + | + | X | + | + | - |
| Strings, 2023 | X | X | + | + | X | X | + | + | X |
| Vicks, 2022 | + | + | + | + | + | X | + | + | + |
| Yoon, 2015 | + | + | + | + | + | + | + | + | + |
| Zethof, 2021 | + | + | + | + | X | X | + | + | - |
| Signorello, 2007 | + | + | + | + | + | + | + | X | + |
| Steinbrecher, 2015 | + | + | + | + | + | + | + | X | + |
| Nyamdorj, 2010 | + | + | + | + | + | + | + | + | + |
| Zhu, 2019 | + | + | + | + | + | + | + | + | + |

D1: Were the criteria for inclusion in the sample clearly defined?
D2: Were the study subjects and the setting described in detail?
D3: Was the exposure measured in a valid and reliable way?
D4: Were objective, standard criteria used for measurement of the condition?
D5: Were confounding factors identified?
D6: Were strategies to deal with confounding factors stated?
D7: Were the outcomes measured in a valid and reliable way?
D8: Was appropriate statistical analysis used?

Judgement: X High; - Unclear; + Low; ? No information

**Figure S1a-b.** Trans-ethnic meta-analysis of the effect of BMI ((a) categorical and (b) per unit increase, HR)

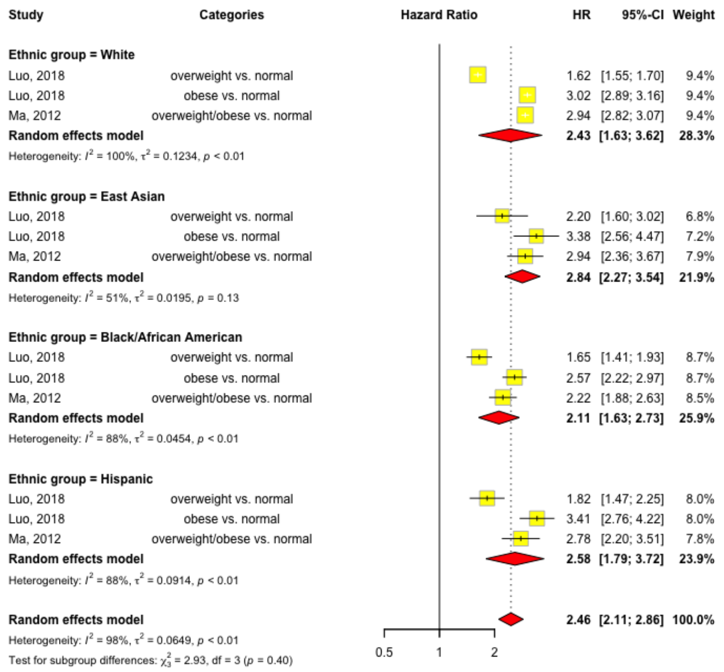


**Figure S1a.**

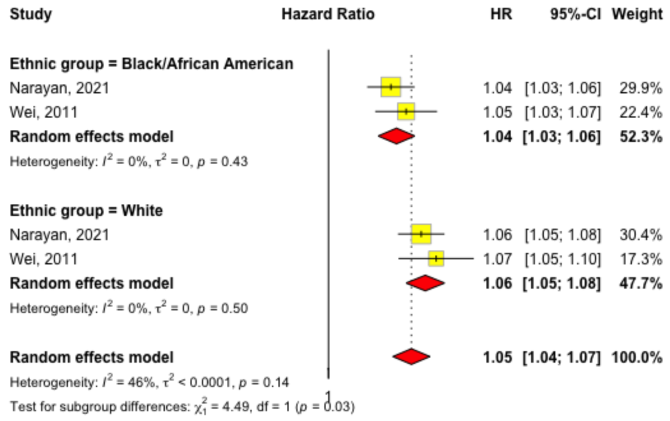


**Figure S1b.**

**Table S8**. Trans-ethnic results of the effect of BMI on T2D from studies not included in the meta-analysis (per SD or unit increase) (Supplementary excel file)

**Table S9.** Trans-ethnic results of the effect of BMI on T2D from studies not included in the meta-analysis (categorical) (Supplementary excel file)

**Table S10**. Trans-ethnic results of the effect of WHR on T2D (Supplementary excel file)

**Table S11**. Trans-ethnic results of the effect of WC on T2D (Supplementary excel file)

**Table S12.** Trans-ethnic results of the effect of WHTR on T2D (Supplementary excel file)

**Table S13**. Trans-ethnic results of the effect of body fat percentage on T2D (Supplementary excel file)

**Figure S2. Funnel plots**

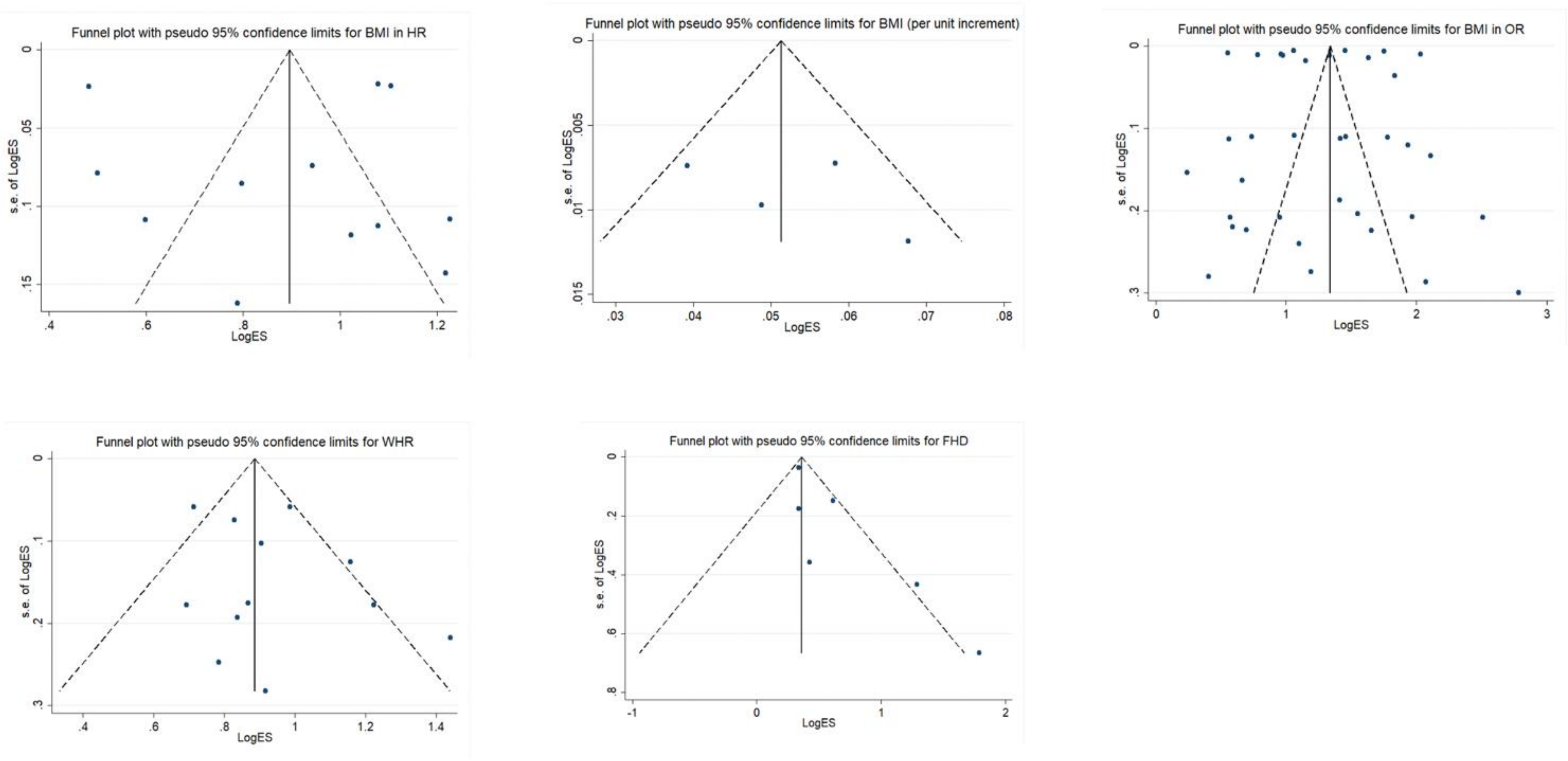

**Figure S3a-c**. Ethnic-specific optimal cut-points of (3a) BMI, (3b) WC and (3c) WHR for T2D (men/women)

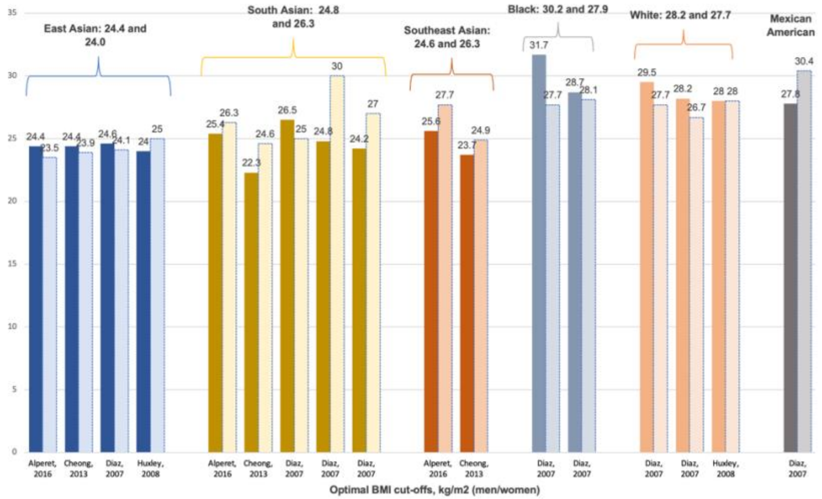


**Figure S3a**

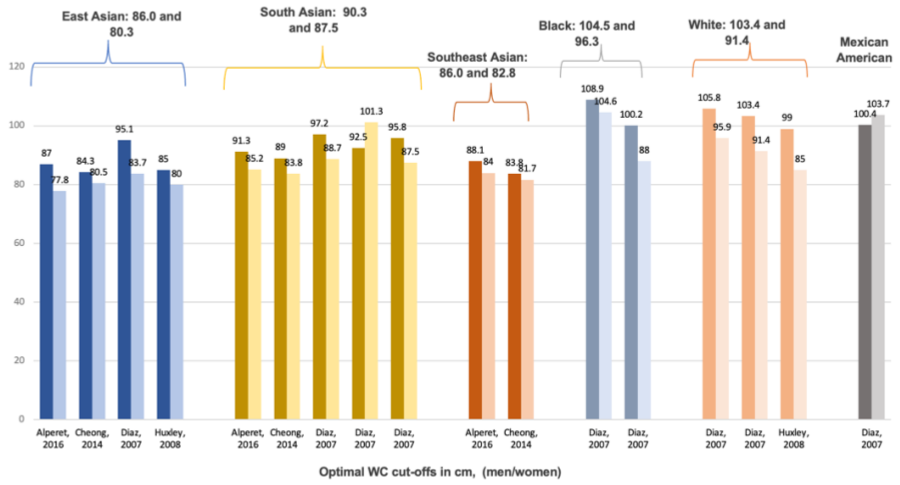


**Figure S3b**

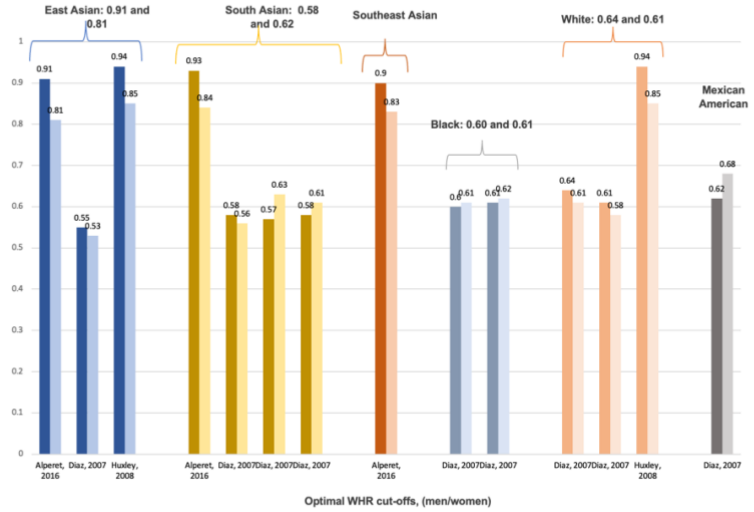


**Figure S3c**

**Table S14.** Trans-ethnic results of the effect of skinfold thickness, the ratio of subscapular to triceps, hand grip strength, WHHR, HC, trunk fat, ABSI, BAI on T2D.

| Phenotype/Risk factor | Study | Results |
|---|---|---|
| **Skinfold thickness (indication of overall obesity)** | Mackay, 2010 | OR (adj.) per SD change in log-transformed sum of skinfold thickness: 2.01 (95% CI 1.40-2.90) for NHW, 1.28 (95%CI 0.85-1.93) for Black/African American and 1.67 (1.18-2.38) for Hispanic group. |
| | Marshall, 1993 | OR per 5mm increase in subscapular skinfold: 1.6 (95%ci 1.4-1.8) for NHW, 1.6 (95%CI 1.4-1.8) for Hispanic group.<br>OR per 5mm increase in triceps skinfold: 1.3 (95%CI 1.1-1.5) for NHW and 1.5 (95%CI 1.3-1.7) for Hispanic group. |
| **The ratio of subscapular to triceps (indication of the ratio of central to peripheral fat)** | Mackay, 2010 | OR (adj.) per SD change in log-transformed measure:<br>2.34 (95% CI 1.51-3.62) for NHW, 2.78 (95% CI 1.65-4.70) for Black/African American, 1.12 (95% CI 0.75-1.67) for Hispanic group. |
| | Marshall, 1993 | OR per 2-unit increase in subscapular to triceps ratio: 1.4 (95%CI 1.3-1.6) and 1.2 (95%CI 1.1-1.3) for Hispanic group. |
| **Hand grip strength** | Ntuk, 2017 | OR for diabetes associated with hand grip strength for men: 1.5 (95%CI 1.4-1.6) for White, 1.6 (95%CI 1.202.0) for Black, 1.4 (1.1-1.7) for South Asian.<br>OR for diabetes associated with hand grip strength for women: 1.2 (95%CI 1.1-1.3) for White, 1.1 (95%CI 0.8-1.3) for Black, 1.6 (95%CI 1.1-2.0) for South Asian. |
| **WHHR (waist-to-hip-height ratio)** | Hardy, 2017b | HR per unit increase (z-score) in WHHR: 1.26 (95%CI 1.17-1.36) for White males, 1.28 (95%CI 1.17-1.40) for Black males, 1.61 (95%CI 1.47-1.77) for White females, 1.44 (95%CI 1.33-1.57) for Black females. |
| **HC (hip circumference)** | Mackay, 2010 | OR (adj.) per SD change in log-transformed measure: 1.75 (95%ci 1.31-2.23) for NHW, 1.05 (95%CI 0.74-1.49) for Black/African American, 1.45 (95% CI 1.09-1.93) for Hispanic group. |
| | Steinbrecher, 2015 | OR per SD increase in HC among men: 1.48 (1.38-1.61) for White, 1.28 (95%CI 1.17-1.41) for Native Hawaiian, 1.48 (95%CI 1.38-1.59) for Japanese American.<br>OR per SD increase in HC among men: 1.67 (95%CI 1.51-1.84) for White, 1.42 (95%CI 1.31-1.54) for Native Hawaiian, 1.69 (95%CI 1.56-1.83) for Japanese American. |
| **ABSI (a body shape index)** | Hardy, 2017a | OR per unit increase (z-score) in ABSI: 1.18 (95%CI 1.02-1.37) for White males, 1.34 (95%CI 1.13-1.60) for Black males, 1.47 (95%CI |

| | | |
|---|---|---|
| | | 1.34-1.61) for White females, 1.44 (95%CI 1.31-1.58) for Black females. |
| | Hardy, 2017b | HR per unit increase (z-score) in ABSI: 1.00 (95%CI 0.94-1.06) for White males, 1.09 (95%CI 0.99-1.19) for Black males, 1.15 (95%CI 1.08-1.22) for White females, 1.19 (95%CI 1.09-1.29) for Black females. |
| **BAI (body adiposity index)** | Hardy, 2017a | OR per unit increase (z-score) in BAI: 1.99 (95%CI 1.72-2.30) for White males, 1.75 (95%CI 1.46-2.11) for Black males, 1.76 (95%CI 1.60-1.92) for White females, 1.28 (95%CI 1.17-1.41) for Black females. |
| | Hardy, 2017b | HR per unit increase (z-score) in BAI: 1.43 (95%CI 1.36-1.49) for White males, 1.35 (95%CI 1.25-1.45) for Black males, 1.54 (95%CI 1.47-1.62) for White females, 1.17 (95%CI 1.12-1.23) for Black females. |
| **Body height** | Jenum, 2005 | OR per one-unit increase (cm) for women: 0.45 (95%CI 0.43-0.47) for White, 1.18 (95%CI 1.08-1.30) for South Asian.<br>OR per one-unit increase (cm) for men: 0.70 (95%CI 0.67-0.73) for White, 0.85 (95%CI 0.79-0.93) for South Asian. |
| **Trunk fat, trunk-to-leg ratio** | Luo, 2019 | HR per unit increase in trunk fat: 1.60 (95%CI 1.51-1.69) for White, 1.40 (95%CI 1.25-1.58) for Black, 1.81 (95%CI 1.47-2.23) for Hispanic.<br>HR per unit increase in trunk-to-leg fat ratio: 1.52 (95%CI 1.44-1.60) for White, 1.69 (95%CI 1.49-2.20) for Black, 1.81 (95%CI 1.49-2.20) for Hispanic. |
| **Combination of anthropometric measures** | Steinbrecher, 2015 | OR per SD increase in WC+BMI for men: 1.60 (95%CI 1.40-1.83) for White, 1.32 (95%CI 1.14-1.54) for Native Hawaiian, 1.59 (95%CI 1.42-1.77) for Japanese American.<br>OR per SD increase in WC+BMI for women: 1.31 (95%CI 1.12-1.53) for White, 1.23 (95%CI 1.08-1.40) for Native Hawaiian, 1.53 (95%CI 1.37-1.71) for Japanese American.<br><br>OR per SD increase in HC+BMI for men: 1.84 (95%CI 1.62-2.09) for White, 1.44 (95%CI 1.24-1.68) for Native Hawaiian, 1.68 (95%CI 1.51-1.86) for Japanese American.<br>OR per SD increase in HC+BMI for women: 1.99 (95%CI 1.66-2.38) for White, 1.51 (95%CI 1.30-1.76) for Native Hawaiian, 2.16 (95%CI 1.89-2.45) for Japanese American.<br><br>OR per SD increase in WHR+BMI for men: 1.70 (95%CI 1.55-1.85) for White, 1.37 (95%CI 1.24-1.51) for Native Hawaiian, 1.65 (95%CI 1.53-1.78) for Japanese American. |

| | | |
|---|---|---|
| | | OR per SD increase in WHR+BMI for women: 1.72 (95%CI 1.55-1.90) for White, 1.47 (95%CI 1.35-1.59) for Native Hawaiian, 1.81 (95%CI 1.67-1.96) for Japanese American.<br><br>OR per SD increase in WHTR+BMI for men: 1.47 (95%CI 1.28-1.69) for White, 1.30 (95%CI 1.12-1.52) for Native Hawaiian, 1.66 (95%CI 1.48-1.87) for Japanese American.<br>OR per SD increase in WHTR+BMI for women: 1.21 (95%CI 1.03-1.43) for White, 1.26 (95%CI 1.10-1.44) for Native Hawaiian, 1.54 (95%CI 1.37-1.73) for Japanese American. |
| | De Koning, 2010 | OR per SD increase in WC+BMI: the stronger effect in Europeans and Aboriginals. Not significant among Black, Hispanic and South Asians.<br>OR per SD increase in WHR+BMI: the strongest effect among Aboriginals, followed by Europeans, Hispanic and South Asians. Not significant among Black ethnic group. |

**Table S15.** Trans-ethnic results of the effect of women-health-associated factors on T2D.

| Phenotype/Risk factor | Study | Results |
|---|---|---|
| **Index of parity** | Almahmeed, 2017 | *Women of Chinese/East Asian* ethnicity (ref. 1): adj.HR 0.90 (95%CI 0.76-1.06) for 2 deliveries, 1.13 (95%CI 0.89-1.43) for 3-4 deliveries, 4.59 (95%CI 2.36-8.92) for ≥5 deliveries.<br><br>*Women of South Asian ethnicity* (ref.1): 0.93 (95%CI 0.81-1.08) for 2 deliveries, 1.03 (95%CI 0.87-1.22) for3-4 deliveries, 1.23 (95%CI 0.81-1.87) for ≥5 deliveries.<br><br>*Women of White ethnicity* (ref. 1): 0.89 (95%CI 0.85-0.93) for 2 deliveries, 1.13 (95%CI 1.08-1.18) for 3-4 deliveries, 1.52 (95%CI 1.39-1.66) for ≥5 deliveries.<br>P for interaction test=0.02. |
| **Birth weight** | Ryckman, 2014 | Birth weight category (ref. 6-7.9 lbs).<br>*White:* OR 1.43 (95%CI 1.25-1.62) for <6lbs., 0.99 (95%CI 0.90-1.10) for 8-9.9 lbs., 1.07 (95%CI 0.88-1.31) for ≥10 lbs. |

| | | |
|---|---|---|
| | | *Black:* OR 1.31 (95%CI 1.04-1.64) for <6 lbs., 1.12 (95%CI 0.91-2.66) for 8-9.9 lbs., 1.44 (95%CI 0.95-2.19) for ≥10 lbs.<br><br>*Asian:* OR 0.87 (95%CI 0.53-2.02) for <6 lbs., 1.56 (95%CI 0.92-2.66) for 8-9.9 lbs., 2.77 (95%CI 0.91-8.43) for ≥10 lbs.<br><br>*Hispanic:* 1.39 (95%CI 0.96-2.02) for <6 lbs., 0.95 (95%CI 0.66-1.37) for 8-9.9 lbs., 0.43 (95%CI 0.13-1.38) for ≥ 10 lbs. |
| **Breastfeeding** | Mayer-Davis, 2008 | *Black/African American women*: OR 0.65 (95%CI 0.24-1.77).<br>*Hispanic women*: OR 0.19 (95%CI 0.05-0.76).<br>*White women*: OR 0.18 (95%CI 0.07-0.49).<br>p-value 0.17 for interaction. Unadjusted model. |
| **Age at menarche** | Dreyfus, 2012 | Adjusted OR: 1.41 (95%CI 1.05-1.89) for White women, 0.94 (95%CI 0.68-1.30) for Black/African American women (p=0.09 for interaction).<br>Adjusted HR: 1.43 (95%CI 1.08-1.89) for White women, 1.20 (95%CI 0.87-1.67) for Black/African American women (p=0.52 for interaction). |

**Figure S4**. Ethnic-stratified meta-analysis results of the effect of FHD on T2D.

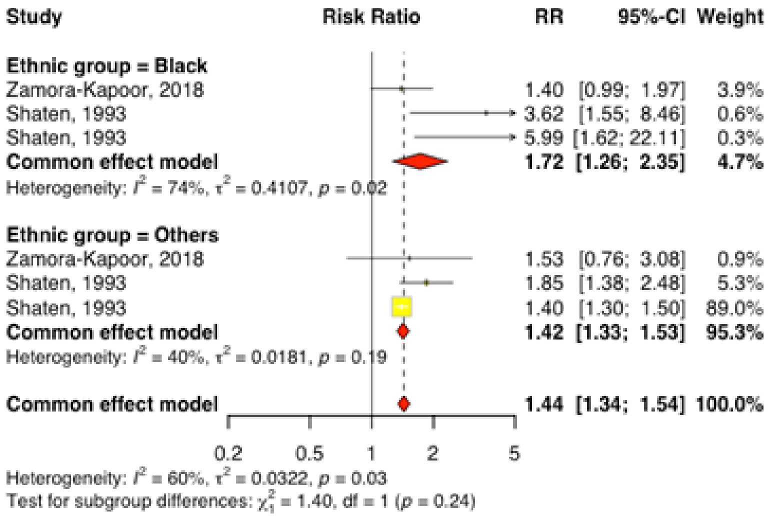


**Table S16**. Trans-ethnic results of the effect of family history of diabetes on T2D from studies not included in meta-analysis (Supplementary excel file)
